# Trending Drugs Combination to Target Leukemia associated Proteins/Genes: using Graph Neural Networks under the RAIN Protocol

**DOI:** 10.1101/2023.08.17.23294228

**Authors:** Mahnaz Boush, Ali A. Kiaei, Hossein Mahboubi

## Abstract

**Background:** Leukemia, a cancer impacting blood-forming tissues such as bone marrow and the lymphatic system, presents in various forms, affecting children and adults differently. The therapeutic approach is complex and depends on the specific leukemia type. Effective management is crucial as it disrupts normal blood cell production, increasing infection susceptibility. Treatments like chemotherapy can further weaken immunity. Thus, a patient’s healthcare plan should focus on comfort, reducing chemotherapy side effects, protecting veins, addressing complications, and offering educational and emotional support.

**Method:** This article reviews studies on the combined use of drugs for treating leukemia. Employing a mix of medicines might decrease the chances of tumor resistance. Starting multiple drugs concurrently allows for immediate application during disease onset, avoiding delays. Initial chemotherapy uses a drug combination to eliminate maximum leukemia cells and restore normal blood counts. Afterwards, intensification chemotherapy targets any residual, undetectable leukemia cells in the blood or bone marrow. To recommend a drug combination to treat/manage Leukemia, under first step of RAIN protocol, we have searched articles including related trend drugs using Natural Language Processing. In the second step, we have employed Graph Neural Network to pass information between these trending drugs and genes that act as potential targets for Leukemia.

**Result:** As a result, the Graph Neural network recommends combining Tretinoin, Asparaginase, and Cytarabine. The network meta-analysis confirmed the effectiveness of these drugs on associated genes.

**Conclusion:** The p-value between leukemia and the scenario that includes combinations of the mentioned drugs is almost zero, indicating an improvement in leukemia treatment. Reviews of clinical trials on these medications support this claim.

**Highlights:**

- Combined drugs that make p-value between Leukemia and target proteins/genes close to 1
- Using Graph Neural network to recommend drug combination
- A Network meta-analysis to measure the comparative efficacy
- Considered drug interactions

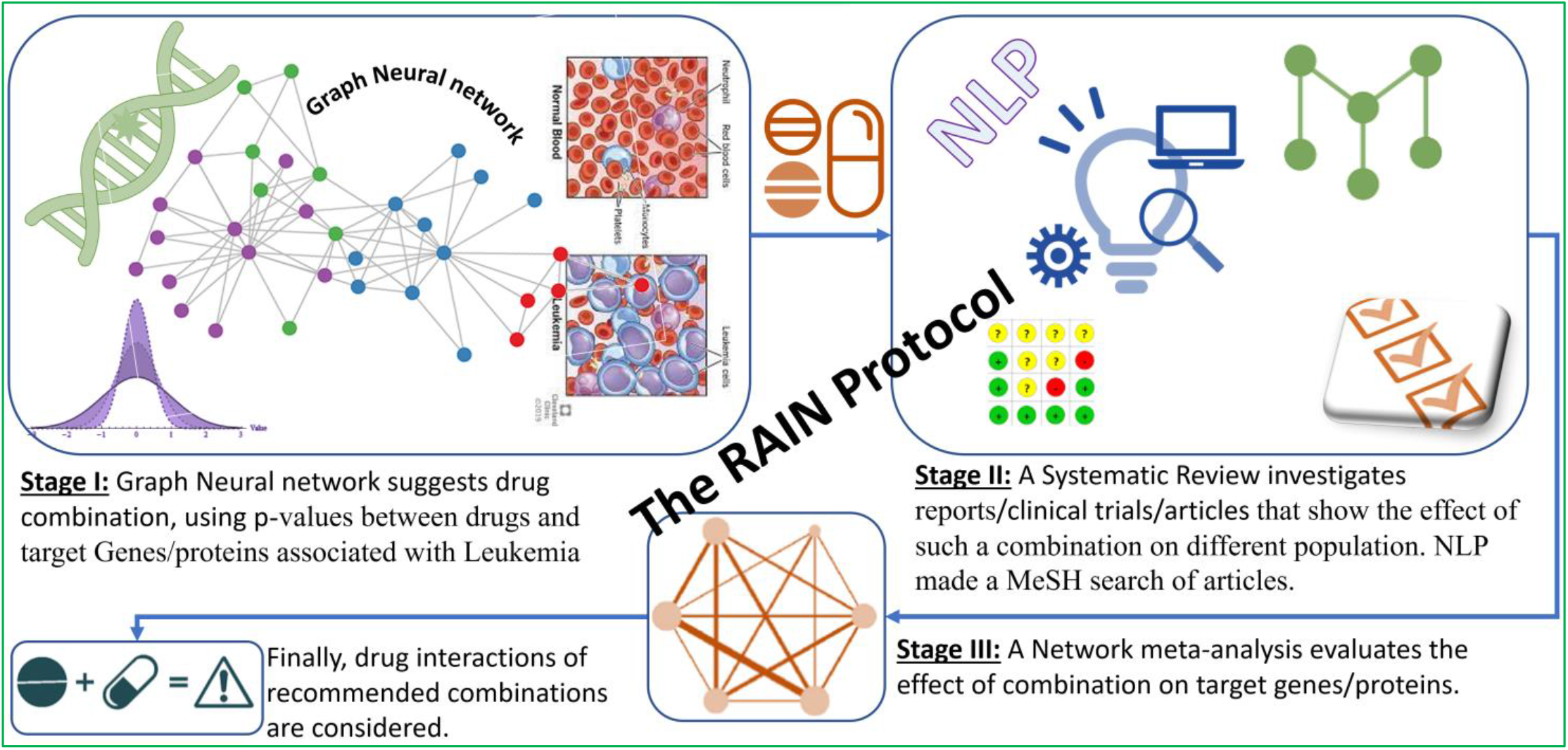

## INTRODUCTION

The introduction of the article is divided into two subsections. The first subsection discusses certain genes or proteins that have been identified as potential targets for the treatment of leukemia. The second subsection provides information about drugs that are currently used to treat leukemia. These drugs may target the genes or proteins mentioned in the first subsection, or they may work through other mechanisms to help fight the disease.

### Associated genes/Proteins

The combination of the PML protein and the RARα protein plays a significant role in the development of Acute Promyelocytic Leukemia (APL). This occurs through the formation of a fusion protein called PML-RARα, which is present in over 95% of APL patients. Due to its high prevalence, this fusion protein becomes a promising target for potential therapeutic interventions. In-depth analysis of PML/RARα’s direct targets has provided valuable insights into the transcriptional abnormalities observed in APL and has paved the way for target-specific treatments. These treatments include all-trans retinoic acid (ATRA) and arsenic trioxide (As2O3), both designed to degrade the PML-RARα protein. ATRA functions by promoting the transcription of RARα-target genes, thereby overcoming the block in cell differentiation. On the other hand, As2O3 induces oxidant stress and directly binds to PML, leading to partial differentiation and cell death in APL cells. Consequently, these treatments prove to be more effective in eradicating leukemia-initiating cells. [1]–[3].

The overexpression of the Leukemia inhibitory factor (LIF) and its receptor (LIFR) is commonly observed in different types of solid cancers, including leukemia. Recent research has identified the LIF/LIFR axis as a potential target for cancer treatment. The activation of signaling pathways that promote cancer growth, such as JAK/STAT3 as immediate effectors, and subsequent downstream MAPK, AKT, and mTOR, is facilitated by LIF/LIFR [4]–[6].

Formerly known as MLL, KMT2A is a gene found on chromosome 11q23 responsible for producing the enzyme histone lysine-specific N-methyltransferase 2A. This gene is frequently involved in recurrent translocations in acute myeloid leukemia (AML), acute lymphoblastic leukemia (ALL), and mixed lineage (biphenotypic) leukemia (MLL) [7]–[9].

RUNX1 is frequently impacted by chromosomal and genetic changes in cases of leukemia. Both inherited and acquired genetic abnormalities are commonly found in acute myeloid leukemia (AML), and the presence of RUNX1 mutations often indicates a poor prognosis. Consequently, there is growing interest in targeting RUNX1 as a potential therapy for AML. Researchers are exploring various strategies to combat RUNX1 in AML, including the development of small molecules that specifically target the RUNX1-RUNX1T1 protein. Additionally, tyrosine kinase inhibitors like dasatinib and FLT3 inhibitors are being investigated to counteract mutations that provide growth advantages to leukemia cells. Epigenetic-based therapies are also being tested as potential approaches to targeting RUNX1 in AML [10], [11].

Around one-third of patients diagnosed with acute myeloid leukemia (AML) have mutations in the fms-like tyrosine kinase 3 (FLT3) gene. It is now recommended that molecular testing for FLT3 mutations be conducted immediately upon diagnosis, and targeted treatments should be promptly integrated to achieve deeper remissions and accelerate consideration for allogeneic stem cell transplant (ASCT). The introduction of the multi-kinase FLT3 inhibitor (FLT3i) midostaurin as part of the initial treatment for newly diagnosed AML patients with FLT3 mutations, as well as the use of the more precise and potent FLT3i gilteritinib as standalone therapy for relapsed/refractory (R/R) AML patients with FLT3 mutations, has significantly improved patient outcomes [12], [13].

Chronic myelogenous leukemia (CML) is a type of blood cancer that arises from a versatile stem cell in the bone marrow and is consistently associated with a specific genetic abnormality known as the BCR-ABL1 fusion gene. This genetic anomaly occurs when a specific gene on chromosome 9, called ABL1, translocates to a region of the BCR gene on chromosome 22. As a result, the BCR-ABL1 fusion gene leads to abnormal activity of a protein called kinase and uncontrolled growth of cells, which are key characteristics of CML. Fortunately, the development of medications known as tyrosine kinase inhibitors (TKIs) that target the BCR-ABL oncoprotein has greatly improved the management of CML [14]–[16].

BCR-ABL, a gene characterized by its abnormal presence in chronic myeloid leukemia (CML) cells, instigates uncontrolled proliferation and replication of these cells. Specifically classified as a tyrosine kinase, the BCR-ABL protein plays a pivotal role in this process. Consequently, tyrosine kinase inhibitors (TKIs) that target BCR-ABL have emerged as the prevailing therapeutic approach for managing CML [17]–[19].

CD19 is a protein commonly present on B-cell acute lymphoblastic leukemia (B-ALL) cells. Remarkable success has been observed in the treatment of B-ALL through the use of Chimeric antigen receptor (CAR) T-cells targeted at CD19. This approach has resulted in complete elimination of the disease in as many as 90% of patients with relapsed or refractory B-ALL [20]–[22].

CD34, a marker for hematopoietic stem cells, has been linked to a less favorable prognosis among individuals diagnosed with myelodysplastic syndromes (MDS) and acute myeloid leukemia (AML). Studies have demonstrated that CD34+CD38-cells possess the ability to initiate AML and B-ALL in individuals with compromised immune systems [23]–[25].

The MECOM gene, also referred to as the MDS1 and EVI1 Complex Locus, is situated on chromosome 3q26.2. This particular gene has undergone extensive research due to its crucial involvement in the maintenance and replication of normal hematopoietic stem cells. However, when it is expressed abnormally, it can act as an oncogene. Translocations that involve the MECOM gene at 3q26.2 have been thoroughly documented and characterized in various myeloid disorders, such as acute myeloid leukemia (AML), myelodysplastic syndromes (MDS), and chronic myelogenous leukemia (CML). These translocations have been associated with a dismal prognosis [26]–[28].

CSF3R functions as the receptor responsible for colony-stimulating factor 3 (CSF3), a critical factor involved in the growth and differentiation of granulocytes. The presence of CSF3R mutations has been observed in individuals suffering from severe congenital neutropenia, a condition that can progress to acute myeloid leukemia (AML) [29], [30].

ZBTB16, a gene associated with leukemia, has recently garnered attention for its involvement in acute myeloid leukemia (AML). In certain instances, a rare genetic rearrangement involving ZBTB16 and the RARA gene has been identified. This rearrangement leads to the creation of a fusion protein called ZBTB16-RARA, which is believed to have oncogenic properties and contribute to the progression of AML [31]–[33].

MEIS1 is a gene that has been linked to the development of leukemia. This gene plays a crucial role as a transcription factor, ensuring the survival of leukemia cells. Consequently, it has emerged as a promising target for potential molecular treatments in cases where leukemias express this particular transcription factor. Studies have revealed that suppressing MEIS1 in leukemia cells leads to a reduction in cell growth, triggers programmed cell death, and slows down the progression of overt leukemia in live organisms [34], [35].

HOXA9, a transcription factor containing a homeodomain, holds significant significance in the expansion of hematopoietic stem cells and is frequently disrupted in cases of acute leukemias. Numerous genetic abnormalities occurring upstream in acute myeloid leukemia (AML) result in the excessive expression of HOXA9, which is a robust indicator of an unfavorable prognosis. While it has been established in several instances that HOXA9 is vital for sustaining leukemic transformation, the precise molecular mechanisms by which it facilitates the development of leukemia are still not fully understood [36].

BCL2, a gene associated with leukemia, has been found to play a significant role in preventing cell death and resistance to chemotherapy in acute myeloid leukemia. This discovery has led researchers from the University of Rochester to investigate the use of BCL2 inhibitors as a potential treatment for leukemia stem cells. These inhibitors have shown promise in targeting and killing leukemia stem cells that are inactive and have slower metabolic rates, suggesting their potential as novel pharmacological modulators in cancer treatment [37]–[39].

DOT1L, a histone methyltransferase, plays a crucial role in the modification of nucleosomal histone H3 lysine 79 (H3K79) through mono-, di-, or trimethylation (H3K79me1, me2, or me3). Its significance lies in its involvement in MLL fusion-driven leukemogenesis, making it a promising target for the treatment of MLL-rearranged leukemias. Extensive research has been conducted, leading to significant advancements in this area. Additionally, EPZ-5676, a DOT1L inhibitor, has successfully entered clinical trials, further highlighting its potential for therapeutic use [40]–[42].

WT1, a leukemia-associated antigen (LAA), exhibits varying expression levels in leukemic blasts. Consequently, WT1 holds potential as a candidate for therapeutic interventions like adoptive-specific T lymphocyte treatments. The restricted presence of WT1 in adult tissues further implies its suitability as a target for combating leukemia [43], [44].

MYC, a proto-oncogene, plays a significant role in a variety of cancers, such as leukemia and lymphoma. Its abnormal expression in hematological malignancies leads to unregulated cell growth and impedes the process of cellular differentiation. Research conducted on leukemia cells and animal models of lymphoma and leukemia indicates that MYC could serve as a promising target for therapeutic interventions. Numerous therapies targeting MYC have been evaluated in preclinical studies and even tested in clinical trials [45]–[47].

MLL is a prime focus for chromosomal translocations in acute leukemias that have a bleak prognosis. The frequently observed fusion partner of MLL, AF9 (also known as MLLT3), has the ability to directly interact with AF4, DOT1L, BCOR, and CBX8. When the direct binding between BCOR and MLL-AF9 is disrupted, there is a partial differentiation of cells and an increase in proliferation. Remarkably, the elimination of the binding between MLL-AF9 and BCOR completely abolished its ability to cause leukemia in a mouse model [48]–[50].

NPM1, a protein that moves between the nucleus and cytoplasm, is primarily found in the nucleolus and plays various important roles. These include controlling the duplication of centrosomes, the production and export of ribosomes, the assembly of histones, the maintenance of genomic stability, and the response to stress in the nucleolus. Mutations in NPM1 are the most common genetic changes found in acute myeloid leukemia (AML), occurring in approximately 30-35% of adult AML cases and over 50% of AML cases with a normal genetic makeup. It has been suggested that dysfunctional NPM1 may contribute to the development of AML by acting as a protein chaperone that prevents leukemia stem cells from differentiating and by regulating non-coding RNAs. In addition to traditional chemotherapy treatments, NPM1 shows promise as a target for AML therapy and should be further investigated [51], [52].

The MLLT10 gene is frequently rearranged in both acute myeloid leukemia (AML) and acute lymphoblastic leukemia (ALL), particularly in T-lineage ALL (T-ALL), across all age groups. Diagnosing and treating MLLT10 rearranged (MLLT10r) acute leukemia is challenging due to the complex nature of the disease, often presenting with an immature or mixed phenotype, and a lack of consensus on the best treatment approach. Patients with MLLT10r AML or T-ALL and an immature phenotype have a high risk of poor outcomes, but the underlying molecular mechanisms and response to targeted therapies are not well understood. By gaining a better understanding of the genomics of MLLT10r acute leukemia, both in clinical and molecular terms, we can improve the ability to predict prognosis and expedite the development of targeted therapies, ultimately leading to better outcomes for patients [53], [54].

TAL1, also known as T-cell acute leukemia protein 1, plays a crucial role in the development of blood cells and the onset of leukemia. Specifically, TAL1 is essential for the differentiation of red blood cells during hematopoiesis. However, it is typically suppressed in the process of thymopoiesis in humans due to epigenetic factors. When TAL1 is abnormally expressed in T-cells, it becomes a significant contributor to the development of T-cell leukemia, which is considered a potent oncogenic event [55], [56].

IL11 receptor (IL11R) is an ideal candidate for targeted therapy in human leukemia and lymphoma due to its presence on the cell surface. A peptidomimetic prototype called BMTP-11 has been developed, which binds specifically to the membranes of leukemia and lymphoma cells. When bound to the IL11R, BMTP-11 triggers the internalization of the ligand-receptor complex, leading to cell death in a dose-dependent manner. These findings suggest that BMTP-11 and its derivatives have promising potential for the treatment of these types of malignancies [57].

The expression of IL3 receptor (CD123) on CD34+CD38-leukemic stem cells in AML and CML has been confirmed. In pre-clinical AML models, it has proven to be a successful target for therapy. The findings also suggest that the IL3 receptor is highly expressed on CD34+38-Bcr-Abl (+) CML stem cells, presenting a promising and achievable target for intervention. Additionally, DT-IL3 conjugates offer a new and innovative approach for selectively targeting CML stem cells that are highly resistant to treatment [58], [59].

CSF2, also referred to as GM-CSF, is a crucial cytokine that regulates a multitude of cellular processes such as differentiation, proliferation, survival, and activation of white blood cells. The receptor responsible for GM-CSF consists of two subunits: CSF2RA and CSF2RB. Notably, CSF2RB serves as a common beta subunit for the IL3 and IL5 receptors, making it the primary subunit for signaling. On the other hand, CSF2RA primarily acts as a subunit that binds to ligands. Studies have shown that GM-CSF signaling plays a inhibitory role in the development of t (8;21) acute myeloid leukemia (AML) in murine models, while also promoting the differentiation of leukemic blasts into myeloid cells [30], [58].

The gene RARA has been linked to the development of acute myeloid leukemia (AML). Through their research, scientists have identified specific areas of DNA called super enhancers (SE) that play a role in overproducing certain gene products in cells from children with AML. Of particular interest is an SE associated with the RARA gene, which was found in 64% of the AML samples obtained from pediatric patients. Further investigation revealed that AML cells containing this RARA SE were highly responsive to treatment with the drug tamibarotene, both in laboratory settings and in animal models. This treatment not only prolonged survival but also effectively reduced the burden of leukemia [31], [60].

JAK2, a protein tyrosine kinase, plays a crucial role in the signaling of cytokine receptors. Recent discoveries have revealed that mutations in JAK2 are associated with the development of acute lymphoblastic leukemia (ALL) and other blood-related cancers. The presence of JAK2 mutations is strongly linked to the formation, treatment, and prognosis of acute leukemia. Moreover, researchers have identified JAK2 and STAT5 as promising targets for therapeutic intervention in leukemic stem cells found in chronic myeloid leukemia (CML) [61]–[63].

CD22, a marker of B-lineage differentiation, has gained significant attention as a promising treatment target for acute lymphoblastic leukemia (ALL). It is highly expressed on the surface of B-precursor ALL cells, making it an ideal candidate for immunotherapy. Importantly, CD22 is found on both immature and mature B cells, while being absent on haemopoietic stem cells. This unique feature positions CD22 as an excellent therapeutic option for patients with relapsed or chemotherapy-refractory ALL [64], [65].

RUNX1 is a commonly affected gene in leukemia, with various changes occurring in its chromosomal and genetic structure. These alterations include rearrangements involving RUNX1 or CBFβ, point mutations in RUNX1, and amplification of the gene. In particular, mutations in RUNX1 are frequently associated with acute myeloid leukemia (AML) and are linked to a poor prognosis. As a result, RUNX1 has emerged as a promising target for therapeutic interventions. Several potential approaches are being explored, including the development of small molecules that can target the RUNX1-RUNX1T1 protein, the use of tyrosine kinase inhibitors like dasatinib and FLT3 inhibitors to counteract mutations that give leukemic cells a growth advantage, and the investigation of epigenetic therapies [66].

BMI1, a gene belonging to the Polycomb-group, is situated at 10p12.2 and plays a significant role in the development of various tumors. It is frequently affected by abnormal changes in chromosomes in B-cell leukemia/lymphoma. Additionally, BMI1 is involved in the preservation of the growth potential of leukemic stem cells (LSCs). By targeting BMI-1, which is substantially elevated in leukemic cells, a substantial reduction in the burden of leukemia was observed [67]–[69].

### Leukemia treatments

Cytarabine plays a crucial role in the treatment of acute myeloid leukemia (AML). It is administered alongside anthracyclines during induction therapy and at higher dosages during consolidation therapy for AML patients. The combination of cytarabine with purine nucleoside analogs, like fludarabine and cladribine, has been extensively studied for the treatment of relapsed or refractory AML patients. However, in recent years, there has been a significant shift towards the use of targeted therapies that are both novel and effective. These therapies include inhibitors that target mutant FMS-like tyrosine kinase 3 (FLT3) and isocitrate dehydrogenase (IDH), as well as venetoclax, a B-cell lymphoma 2 inhibitor, and glasdegib, a hedgehog pathway inhibitor. In older patients, a combination of a hypomethylating agent or low-dose cytarabine with venetoclax has shown promising results, achieving response rates similar to standard induction regimens in similar populations. However, this combination therapy may come with reduced toxicity and lower risk of early mortality [70]–[72].

For over 40 years, daunorubicin, an anthracycline drug, has been widely used alongside cytarabine as a standard induction therapy for adult patients with acute myeloid leukemia (AML). This treatment approach, commonly referred to as the “3+7 regimen,” involves administering daunorubicin for three days followed by seven days of cytarabine. Recently, a phase 2 trial examined the effectiveness and safety of combining venetoclax with the 3+7 regimen in adult AML patients. The results demonstrated an impressive composite complete remission rate of 91% after just one cycle of treatment [73]–[75].

Mercaptopurine, a potent chemotherapy medication, is primarily utilized in the treatment of acute lymphoblastic leukemia (ALL), a type of cancer affecting the white blood cells. Additionally, it can be employed to address acute myeloid leukemia (AML), as well as the rare form of AML known as acute promyelocytic leukemia. This medication acts as a pro-drug, resembling the structure of purine analogues, and disrupts the synthesis and recycling of nucleotides within the body. For the successful eradication of ALL, it is crucial to undergo maintenance therapy (MT) involving the administration of oral methotrexate (MTX) and 6-mercaptopurine (6-MP). The primary method by which this drug exerts its cytotoxic effects is through the integration of thioguanine nucleotides (TGNs) into the DNA molecule. This process may be further intensified due to the inhibition of the production of new purines by other metabolites derived from MTX/6-MP [76]–[78].

Vincristine, an effective chemotherapy medication, is frequently administered alongside other drugs for the treatment of acute lymphoblastic leukemia (ALL). This compound works by interfering with the normal functioning of microtubules, thereby halting the cell cycle. In order to assess the possibility of discontinuing pulse therapy with vincristine and dexamethasone beyond one year of treatment for childhood ALL, a study was conducted to determine if this approach would have any negative impact on the outcome of any risk subgroup within this population [79]–[82].

Methotrexate is a chemotherapy drug commonly used to treat acute lymphoblastic leukemia (ALL). This medication works by depleting reduced folates and inhibiting key steps in nucleotide synthesis, which are necessary for the production of thymidine and purine. Maintenance therapy, consisting of oral methotrexate and 6-mercaptopurine, is crucial for successfully curing ALL. These drugs disrupt nucleotide synthesis and salvage pathways, and their primary method of killing cancer cells involves incorporating thioguanine nucleotides into DNA. This process can be further enhanced by the inhibition of de novo purine synthesis by other metabolites of methotrexate and 6-mercaptopurine [83]–[87].

Busulfan, a chemotherapy medication, is utilized alongside other drugs to prepare patients with leukemia for allogeneic marrow transplantation. For individuals with acute myeloid leukemia or myelodysplastic syndrome who are deemed unfit for intense conditioning treatments, a widely accepted reduced-intensity conditioning regimen involves a lower dosage of intravenous busulfan combined with the purine analogue fludarabine [88]–[92].

Cyclophosphamide, a chemotherapy medication, has shown potential in enhancing the effectiveness of treatment for patients with acute lymphoblastic leukemia (ALL). Despite its promising results, there remains a lack of clarity when it comes to assessing the overall efficacy and safety of this drug for ALL patients [93], [94].

Doxorubicin has been proven to be a highly effective chemotherapy medication in the treatment of acute lymphoblastic leukemia (ALL) in children. However, recent developments in the form of bispecific antibodies (BsAbs) have further enhanced its effectiveness by improving its ability to target leukemia cell lines and patient samples. This is particularly beneficial as these samples often consist of varying immunophenotypes and represent high-risk subtypes of childhood leukemia. Moreover, the use of a clinically approved and low-toxic PEGylated liposomal formulation of doxorubicin, known as Caelyx, in combination with BsAbs has shown promising results in increasing the cytotoxic activity against these leukemia cells [95]–[97].

Asparaginase, an enzyme known for its ability to reduce the levels of the amino acid L-asparagine in the bloodstream, plays a crucial role in the treatment of acute lymphoblastic leukemia (ALL). By depriving leukemic cells of this amino acid, which they are unable to produce themselves, asparaginase effectively induces cell death. Moreover, this enzyme has the unique capability of reconfiguring the metabolic and energy-producing pathways within leukemia cells, thereby triggering both anti-leukemic and pro-survival mechanisms [98]–[100].

Tretinoin, also known as all-trans-retinoic acid (ATRA), is a medication utilized to induce remission in individuals diagnosed with acute promyelocytic leukemia (APL) who possess a specific gene mutation known as the t(15;17) translocation, resulting in the formation of the PML::RARα fusion gene. This medication is not prescribed for maintenance therapy. Researchers have examined the combination of arsenic trioxide and tretinoin (AsO/ATRA) as a potential treatment approach for APL. For individuals newly diagnosed with APL and exhibiting low-risk disease (with a white blood cell count [WBC] of ≤10,000/mcL), AsO/ATRA is the preferred induction regimen. Additionally, patients with high-risk disease (WBC >10,000/mcL) who are unable to tolerate anthracyclines are advised to undergo the AsO/ATRA induction regimen [101]–[103].

Imatinib, a targeted inhibitor of BCR-ABL1 kinase, has significantly enhanced the outlook for chronic myeloid leukemia (CML) patients. Examining the efficacy and safety data from over a decade of follow-up on CML patients treated with imatinib as their initial therapy, it is evident that imatinib’s effectiveness remains consistent over time. Furthermore, long-term use of imatinib does not lead to any concerning cumulative or delayed toxic effects [104], [105].

Idarubicin, an anthracycline medication, demonstrates remarkable efficacy in combating acute myeloid leukemia (AML). Its oral administration capability allows for the development of anti-leukemic treatment plans that can be conveniently administered via oral intake. This feature proves invaluable, particularly for elderly AML patients who are deemed unfit for conventional intensive therapies [106]–[108].

Etoposide, a potent anticancer medication, has demonstrated effectiveness as a standalone treatment for acute myeloid leukemia (AML). Nevertheless, the administration of high dosages or prolonged use of etoposide may lead to the development of therapy-related leukemia [109]–[111].

Thioguanine, a potent anti-cancer medication primarily utilized in leukemia treatment, is now being investigated through a study called Thiopurine Enhanced ALL Maintenance (TEAM). The objective of this study is to assess the potential enhancement in disease-free survival by incorporating a minute quantity of 6-thioguanine into the maintenance therapy based on 6-mercaptopurine/methotrexate. The study focuses on both pediatric and adult patients, ranging from 0 to 45 years of age, who have recently been diagnosed with B-cell precursor or T-cell acute lymphoblastic leukemia. These patients are being treated in accordance with the intermediate risk-high group of the ALLTogether1 protocol [86], [112]–[114].

The combination of arsenic trioxide and all-trans retinoic acid (ATRA) has proven to be highly effective in treating acute promyelocytic leukemia (APL), leading to significant advancements in patient outcomes. As a result, APL has emerged as the most easily treatable form of acute myeloid leukemia [101], [102], [115], [116].

Fludarabine, an antineoplastic drug, is widely employed in the management of hematological malignancies, specifically chronic lymphocytic leukemia (CLL) and indolent B-cell lymphoma. Due to its ability to suppress the immune system, fludarabine has been incorporated into reduced intensity conditioning regimens [27], [117]–[119].

Amsacrine, an antineoplastic agent, has shown promising results in treating acute leukemias. However, it is important to note that using Amsacrine-based induction therapy in patients with cardiac comorbidities should not be considered as an alternative to the standard induction treatment for acute myeloid leukemia (AML) [120]–[123].

Mitoxantrone is a potent therapeutic agent widely employed in the management of acute leukemia. In various clinical investigations involving individuals with refractory and relapsed acute myeloid leukemia (AML), a combination of Mitoxantrone and etoposide has been utilized, yielding notable rates of complete remission (CR) ranging from 16% to 61% [109], [124], [125].

Cytosine arabinoside, which is commonly referred to as cytarabine, holds significant value as a medication for addressing acute myeloid leukemia (AML). It plays a crucial role in induction therapy when combined with anthracyclines and in consolidation therapy at elevated doses for AML patients³. Although high-dose cytarabine (ranging from 2000 to 3000 mg per square meter of body-surface area) can be toxic, it yields superior rates of relapse-free survival compared to the traditional dosage of 100 to 400 mg per square meter [71], [126], [127].

The effectiveness of Recombinant Interleukin-3 (IL-3) in combating leukemia has been explored through research. Specifically, a new fusion protein called DT388IL3 has been developed for this purpose. DT388IL3 is a combination of the catalytic and translocation domains of diphtheria toxin (DT388) and human interleukin 3 (IL-3), connected by a Met-His linker. In order to assess its potential in treating differentiated human acute myeloid leukemia (AML), DT388IL3 was tested in an in vivo model [128]–[131].

Transplant conditioning stands as a critical component in the treatment procedure for leukemia. Prior to the transplant, medical professionals employ chemotherapy, radiation therapy, or a combination of the two, aiming to eliminate as many cancerous cells as feasible. This phase of treatment not only weakens the immune system to minimize the chances of transplant rejection but also creates space for the introduction of fresh stem cells. Conditioning regimens assume an integral role in the complete eradication of leukemic cells [88], [132]–[134].

Myeloablative conditioning is a preparative regimen employed prior to a stem cell transplant, aimed at achieving sufficient immunosuppression to prevent rejection of the transplanted graft and eliminate the underlying disease. When treating acute lymphoblastic leukemia (ALL), myeloablative conditioning typically involves the use of total-body irradiation (TBI) or busulfan. TBI-based regimens are particularly effective in eradicating leukemia cells in hard-to-reach areas, and as a result, cyclophosphamide and TBI are currently the preferred myeloablative regimen for ALL [132], [135]–[137].

The preparative regimen, also referred to as the conditioning regimen, plays a crucial role in the hematopoietic cell transplant (HCT) procedure. Its primary objectives are twofold: to ensure adequate immunosuppression to prevent the rejection of the transplanted graft and to eliminate the disease for which the transplant is being conducted. There are two main types of preparative regimens: standard-intensity and reduced-intensity. The standard-intensity regimen involves the administration of high doses of chemotherapy, sometimes combined with high doses of radiation, and is commonly known as a myeloablative regimen. On the other hand, the reduced-intensity regimen employs lower doses of chemotherapy, possibly with lower doses of radiation, and is commonly referred to as a non-myeloablative regimen [138].

Anthracyclines, which are chemotherapy drugs derived from various strains of Streptomyces bacteria, enjoy extensive usage in the treatment of a wide range of cancers. These include leukemias, lymphomas, as well as cancers affecting the breast, stomach, uterus, ovary, and lung, among other locations. The mechanism of action for anthracyclines involves the infliction of DNA damage upon cancer cells, ultimately leading to their demise before they can undergo cell division. Numerous variations of anthracycline drugs are employed in chemotherapy, with certain medications exhibiting remarkable efficacy in targeting specific cancer types [139].

Filgrastim is a pharmaceutical drug employed to bolster the production of white blood cells in individuals undergoing chemotherapy for acute myeloid leukemia (AML). Its purpose is twofold: to expedite recovery from fever and to minimize the risk of infection in patients with neutropenia resulting from specific types of chemotherapy, including those prescribed for AML [140].

Prednisolone, a potent steroid, serves a dual purpose in medical applications - combating leukemia cells and alleviating allergic reactions caused by certain chemotherapy medications. Its primary function is to curtail inflammation and modulate the body’s immune response. Prednisone, a popular glucocorticoid, has emerged as the standard treatment for individuals diagnosed with acute lymphoblastic leukemia (ALL). Typically administered for a duration of four consecutive weeks, it is complemented by a combination of other chemotherapy drugs [121], [141], [142].

Tyrosine kinase inhibitors (TKIs) are a class of drugs widely recognized as the standard treatment for chronic myeloid leukemia (CML). These medications function by deactivating the tyrosine kinase, which is produced by the BCR-ABL1 gene present in leukemia cells. By doing so, they effectively halt or slow down the abnormal production of white blood cells in the bone marrow. Notable examples of TKIs employed in the management of CML encompass Imatinib (Gleevec), Dasatinib (Sprycel), Nilotinib (Tasigna), Bosutinib (Bosulif), Ponatinib (Iclusig), and Asciminib (Scemblix) [15], [17], [143]–[145].

Azacitidine, a prescribed medication, serves as an effective remedy for specific leukemia forms, such as acute myeloid leukemia (AML) and myelodysplastic syndromes (MDS). This medication is specifically formulated to impede the excessive growth of leukemia cells while simultaneously promoting the production of healthy and fully functioning cells in the bone marrow. The primary objectives of this therapy encompass elevating blood cell counts, mitigating the susceptibility to infections, reducing the dependency on blood transfusions, and minimizing the risk of bleeding. Notably, Azacitidine is highly recommended as the initial course of treatment for elderly AML patients who are unsuitable candidates for intensive treatment regimens [146]–[148].

Clofarabine, a second-generation purine analog, is a highly effective medication prescribed for the treatment of acute lymphoblastic leukemia (ALL) in children and young adults, specifically those aged between 1 and 21 years, who have undergone at least two unsuccessful treatment methods. Typically administered as a last resort, this potent drug exerts its therapeutic effects by strongly impeding DNA synthesis while exhibiting a favorable pharmacologic profile [149], [150].

Aclarubicin is a pharmaceutical utilized for the therapeutic management of acute non-lymphocytic leukemia, a malignancy affecting the blood and bone marrow. In the context of acute myeloid leukemia (AML), this medication has been administered as an induction therapy for patients who have shown resistance to initial chemotherapy or have experienced a relapse [151], [152].

Dasatinib, a second-generation BCR-ABL1 kinase inhibitor, is an effective medication prescribed for the treatment of specific forms of leukemia, such as chronic myeloid leukemia (CML) and Philadelphia chromosome-positive acute lymphoblastic leukemia (ALL). It is particularly approved as an initial therapy for CML and for adult patients who no longer derive benefits from alternative leukemia medications, such as imatinib (Gleevec), or for those who are unable to tolerate them due to adverse reactions [153]–[155].

Tetradecanoylphorbol Acetate (TPA) is a highly effective tumor promoter frequently utilized in the field of biomedical research to activate the signal transduction enzyme protein kinase C (PKC)². Furthermore, TPA has demonstrated remarkable potential as a powerful inducer of differentiation in human promyelocytic leukemia cells¹. Notably, TPA has proven successful in treating patients with myelocytic leukemia, resulting in therapeutic effects and temporary remission. However, further investigation is required to determine the optimal dosing regimens of TPA and assess whether it can lead to long-lasting or even permanent remissions of myelocytic leukemia [156]–[158].

Venetoclax, a pharmaceutical medication, has obtained official approval for the treatment of newly diagnosed acute myeloid leukemia (AML). Specifically designed for individuals aged 75 years and older, as well as adults who are unable to undergo intensive induction chemotherapy, this drug is administered alongside azacitidine, decitabine, or low-dose cytarabine. Furthermore, Venetoclax has exhibited considerable potential in addressing AML cases characterized by heightened Bcl-2 levels [73], [107], [159].

#### Objectives

Despite the existence of various studies on useful drugs for leukemia, the reasons shown in Figure 1 have led to lack of comprehensive statistical research on the subject. The use of artificial intelligence, on the other hand, has been considered in various medical applications in recent years, from protein folding [160], medical imaging[161], [162], cohort studies[163], until core fundamental changes in neural networks[164].

**Figure 1:**
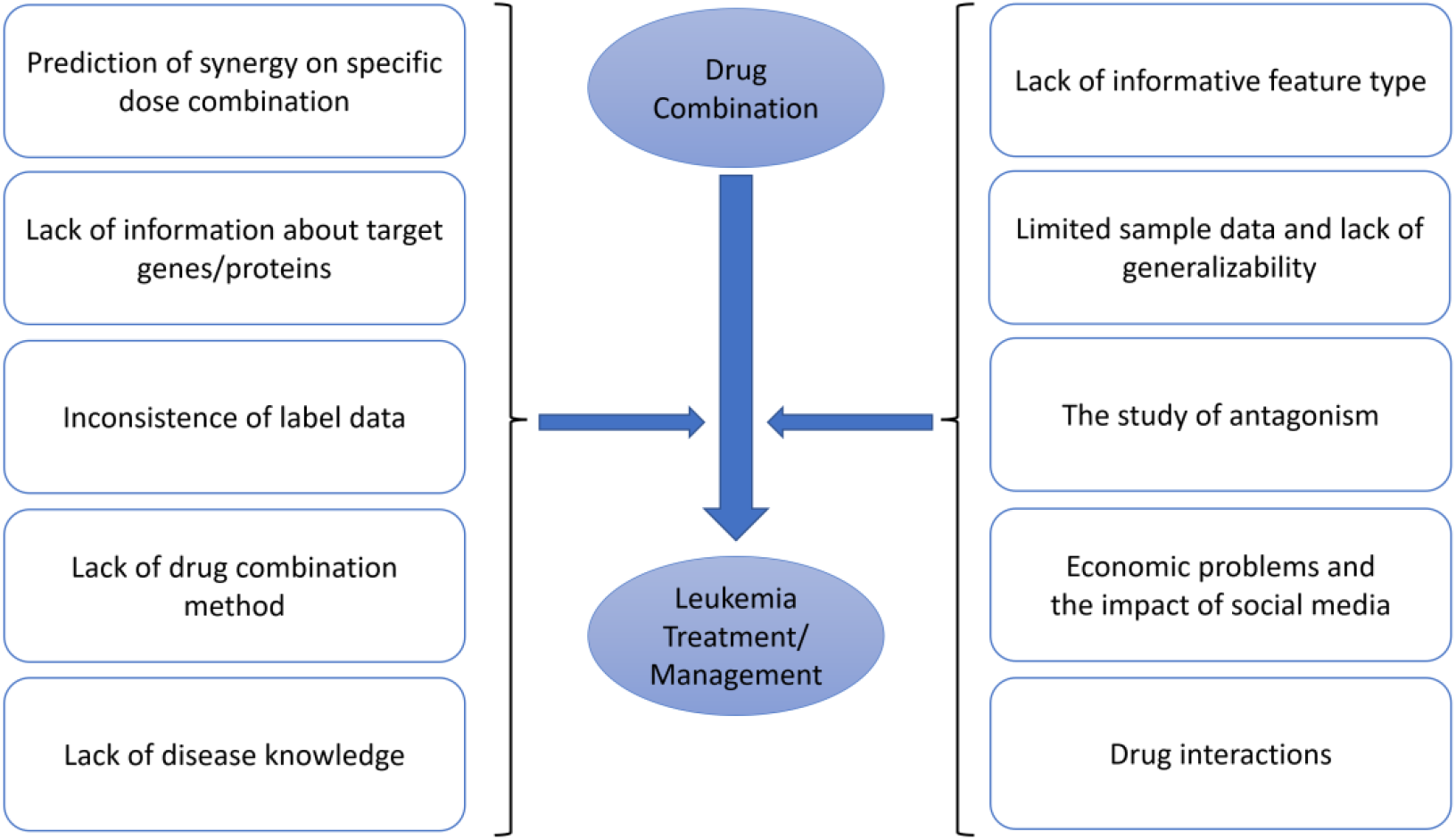
The effects of proposed drug combinations on the management of Leukemia incidents.

The goal of this research is suggesting drug combinations to manage/treat Leukemia using the RAIN protocol, which employs artificial intelligence to recommend drug combinations for managing or treating diseases [165]. The RAIN protocol has been used in recent years to propose medicinal compounds for diseases such as cancers [165]–[170].

## METHOD

The RAIN protocol comprises three distinct stages. Initially, artificial intelligence is employed to propose an optimal drug combination for the treatment and management of a specific disease. Subsequently, a comprehensive analysis is conducted through Natural Language Processing, systematically reviewing recent articles and clinical trials to assess the effectiveness of various permutations of the suggested combination. Figure 2 presents the distribution of articles in each step of the STROBE checklist. Finally, in the third stage, the effectiveness of drugs and their associated human proteins/genes is evaluated using Network meta-analysis.

**Figure 2:**
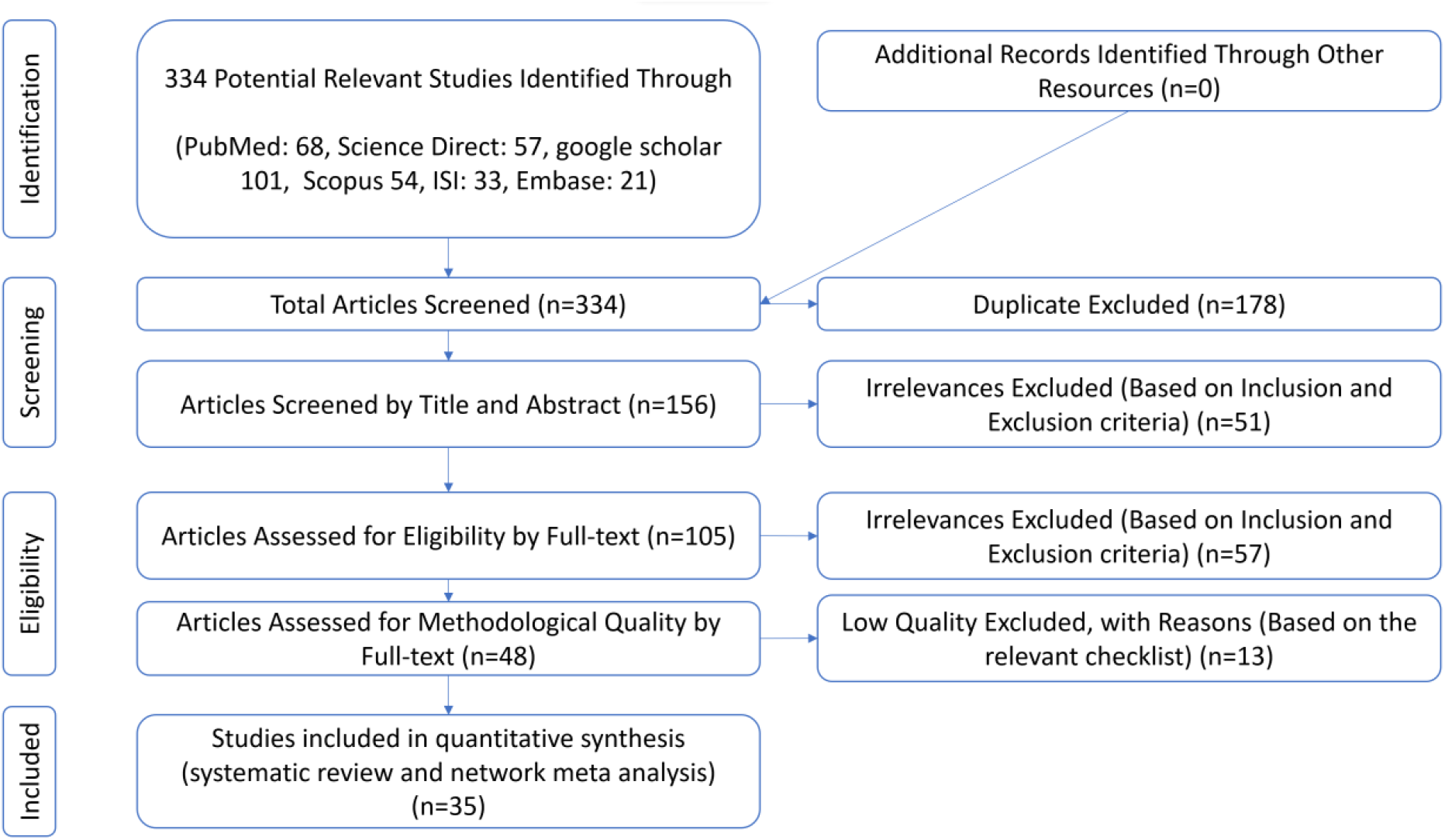
PRISMA (2020) flow diagram indicating the stages of sieving articles in this RAIN protocol.

### Stage I: Graph Neural Network

The RAIN protocol incorporates the GNN (Graph Neural Network) approach to foster collaboration among associations towards a specific goal. This strategy utilizes a network meta-analysis to identify associations with known p-values, which are then transformed into cooperations with limited significance. The proposed model consists of two main steps: the forward and backward steps. During the forward step, the GNN algorithm calculates combined p-values between associations and the target, ultimately selecting the association with the lowest combined p-value. In the subsequent backward step, p-values between interface features and the target are updated based on the chosen associations. The significance of these cooperative associations is determined by multiplying their combined p-values. This iterative process continues until the significance falls below a predetermined threshold. A visual representation of this process can be observed in Figure 3.

**Figure 3:**
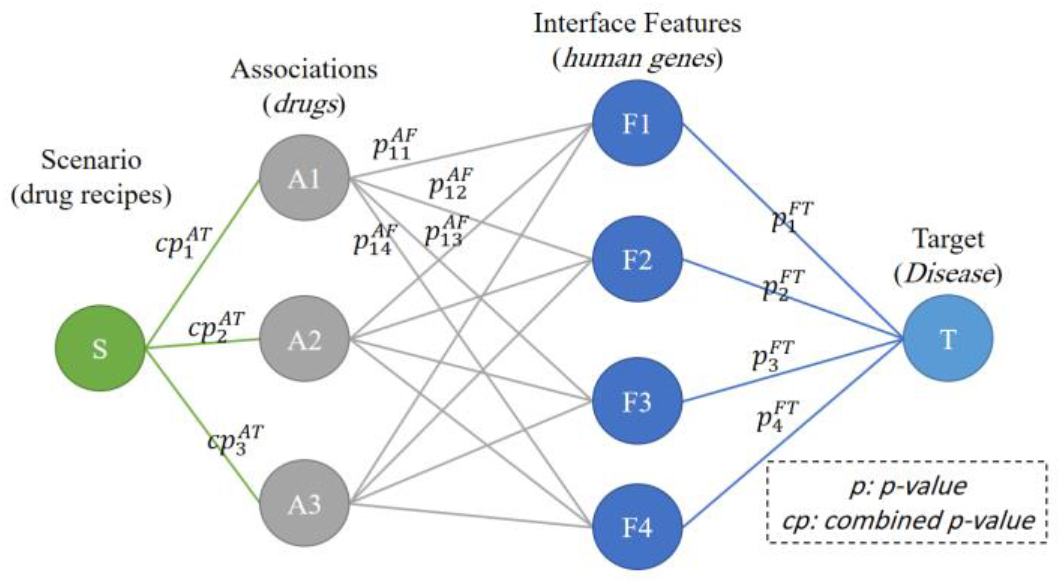
The general structure of the GNN model to suggest an effective drug combination in the management of disease using human proteins/genes as interface features.

### Stage II: A comprehensive Systematic Review

This stage outlines a procedure for validating the results obtained from a GNN model by conducting a systematic evaluation of recommended medications. To carry out this evaluation, a systematic review is performed using various databases including Science Direct, Embase, Scopus, PubMed, Web of Science, and Google Scholar. Instead of manually searching within these databases, a semantic search technique based on Natural Language Processing (NLP) is employed. This technique utilizes MeSH to search for each term individually, resulting in a wider and more precise selection of articles within a shorter timeframe.

#### Information sources

In order to validate the proposed drug combination suggested by our in-house GNN model, we conducted a comprehensive systematic review utilizing NLP techniques. Our review encompassed multiple databases such as Science Direct, Embase, Scopus, PubMed, Web of Science, and Google Scholar, with the aim of identifying relevant studies. The primary objective was to analyze data from a large-scale clinical trial, thereby affirming the efficacy of the suggested drug combination. To accomplish this, we extracted keywords from both the GNN model outputs and the Leukemia subscription.

#### Search strategy

The utilization of a natural language processing method enables the execution of a semantic search across diverse databases, specifically targeting publication titles and abstracts. This advanced technique facilitates the inclusion of MeSH phrases as potential search keywords, as their semantic value proves to be highly valuable in the search process.

#### Study selection

In the initial stage of the process, the elimination of redundant research is prioritized. Subsequently, a comprehensive list of the remaining research titles is compiled during the evaluation phase in order to systematically filter the research. In the screening phase, which is the first step of the systematic review, a thorough assessment of the titles and abstracts of the remaining research is conducted, and certain studies are excluded based on predefined selection criteria.

In the subsequent step, known as the competency evaluation, the full text of the research that survived the screening phase is meticulously reviewed in accordance with the selection criteria, resulting in the elimination of several studies that are not relevant. To ensure objectivity and minimize personal bias in the selection of resources, an expert and a Question-Answering (QA) agent utilizing Natural Language Processing (NLP) independently carry out the research and data extraction process. The expert is required to provide a comprehensive explanation for any study that was not chosen. Meanwhile, the QA agent assigns a score to each article based on specific questions, and articles with the lowest scores are excluded. These questions involve determining the effectiveness of different drugs for the treatment of leukemia, with each drug being substituted in the intelligent system’s output. In cases where there is disagreement between the expert and the QA agent, the expert will review the contentious research.

#### Quality evaluation

In order to assess the quality of remaining publications in a specific research field, a checklist is commonly employed. The STROBE method is frequently utilized to evaluate the quality of observational studies. This checklist is structured into six comprehensive sections, namely title, abstract, introduction, methodology, results, and discussion. It encompasses a total of 32 fields, each of which corresponds to a distinct aspect of a study’s methodology. These fields include elements such as the study’s title, problem statement, objectives, type, population, sampling method, sample size, variable and procedure definitions, data collection methods, statistical analysis techniques, and results. The maximum score attainable during the quality assessment phase using the STROBE checklist is 32. Articles that receive a score of 16 or higher are regarded as being of moderate or high quality.

### Stage III: Network meta-analysis

In the third phase, a network meta-analysis is applied to investigate the impact of potential synthetic drug combinations on human proteins/genes. This approach allows for the evaluation of multiple drugs within a single study, combining both direct and indirect data from randomized controlled trials that link diseases and drugs through proteins/genes. By utilizing proteins/genes as a connecting element in a network, this analysis helps to determine the relative effectiveness of commonly prescribed drugs in real-world clinical settings. The evaluation of each drug’s efficacy is based on input biological data.

## RESULTS

### Stage I: Graph Neural network

The recommended drug combination by the GNN consists of Tretinoin, Asparaginase, and Cytarabine. The significance of this combination is demonstrated in Table 1, which displays the p-values associated with the combination of these drugs. For instance, when considering the p-value between Leukemia and Tretinoin (Scenario 1), the value is 0.01. However, this value decreases significantly to 0.000088 when Asparaginase is added (Scenario 2). Moreover, the p-value further decreases to 3E-5 in the third scenario, indicating that the proposed drug combination has a positive impact on managing the disease.

**Table 1:** p-value between scenarios and Leukemia.

| *Scenario* | *Drug Combinations* | *p-value* |
| --- | --- | --- |
| *S1* | Tretinoin | 1.0193E-02 |
| *S2* | S1 + Asparaginase | 8.7631E-05 |
| *S3* | S2 + Cytarabine | 3.4965E-05 |

Table 2 provides insights into how the p-values between Leukemia and human proteins/genes change under different scenarios. The ‘S0’ column showcases the p-values between Leukemia and the respective affected human proteins/genes. When Tretinoin is introduced (S1 column), the combined p-values are shown. It is interesting to note that in the ‘S3’ column, the p-values between Leukemia and human proteins/genes reach 1, implying a decrease in the significance of the target proteins/genes.

**Table 2:**
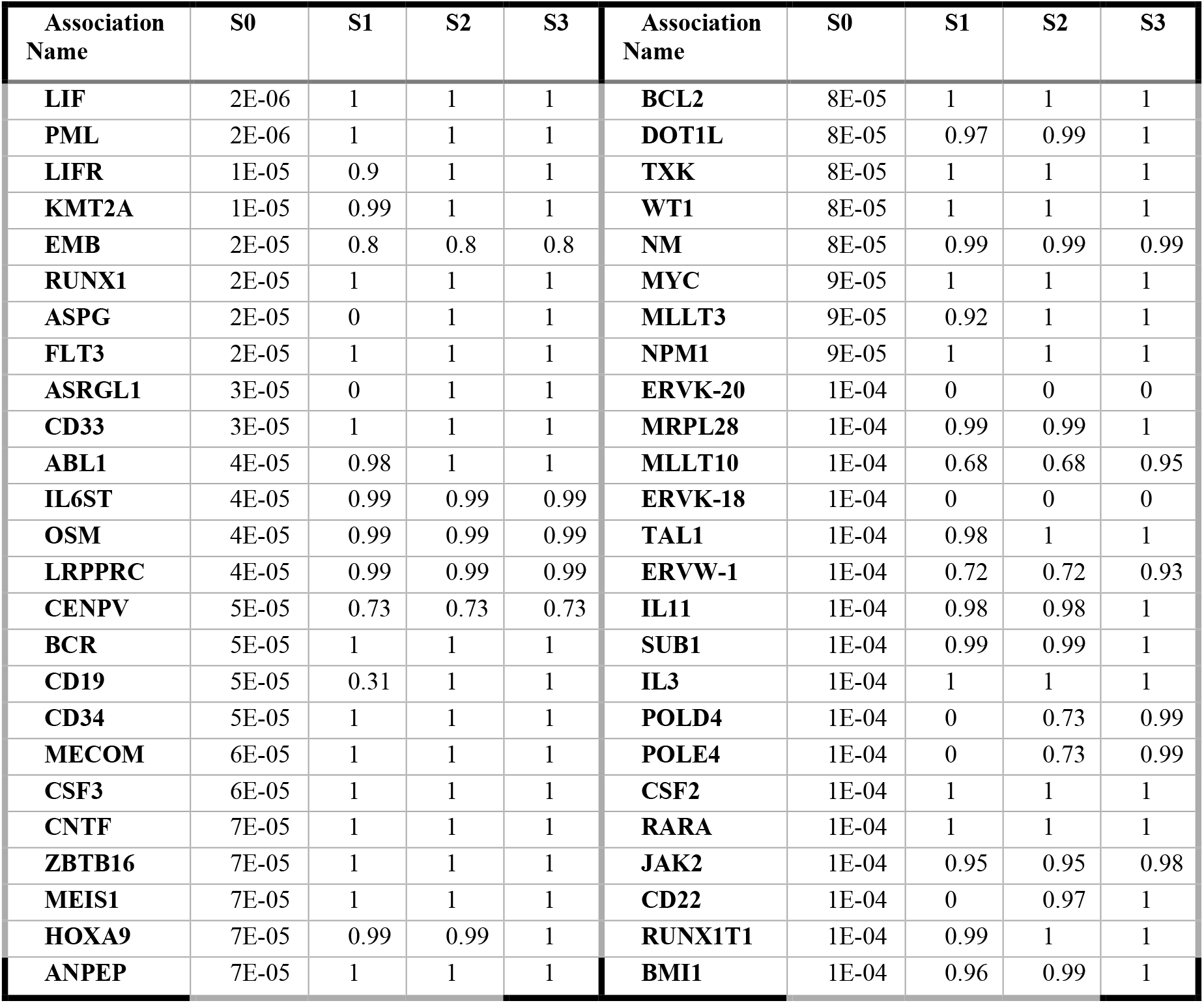
p-values between Leukemia and human proteins/genes after implementing scenarios.

### Stage II: A comprehensive Systematic Review

The impact of the mentioned medications on leukemia treatment was thoroughly investigated at this stage. A rigorous selection process was followed, adhering to the PRISMA principles and the RAIN framework, to gather relevant articles until July 2022. A total of 251 potentially relevant articles were identified and imported into the EndNote reference management system. After removing duplicates (178 articles), the remaining 131 papers were subjected to title and abstract review, using predefined inclusion and exclusion criteria. Consequently, 40 studies were excluded during the screening phase. Moving on to the eligibility evaluation, 120 studies met the criteria and their full texts were thoroughly reviewed. Subsequently, 47 studies were eliminated based on their adherence to the inclusion and exclusion criteria. In the quality evaluation stage, the remaining 16 studies were assessed using the STROBE checklist scores and methodological quality, resulting in the exclusion of 15 studies due to poor quality. Ultimately, 35 cross-sectional studies were included in the final analysis. The full texts of these articles were meticulously examined, and each article was scored using the STROBE checklist (refer to Figure 3). For further details and characteristics, please refer to Table 3 [171]–[205]. Additionally, the structures of the drugs used in the studies are displayed in Figure 4, while Table 4 provides an overview of the drug properties [171]–[205].

**Table 3:**
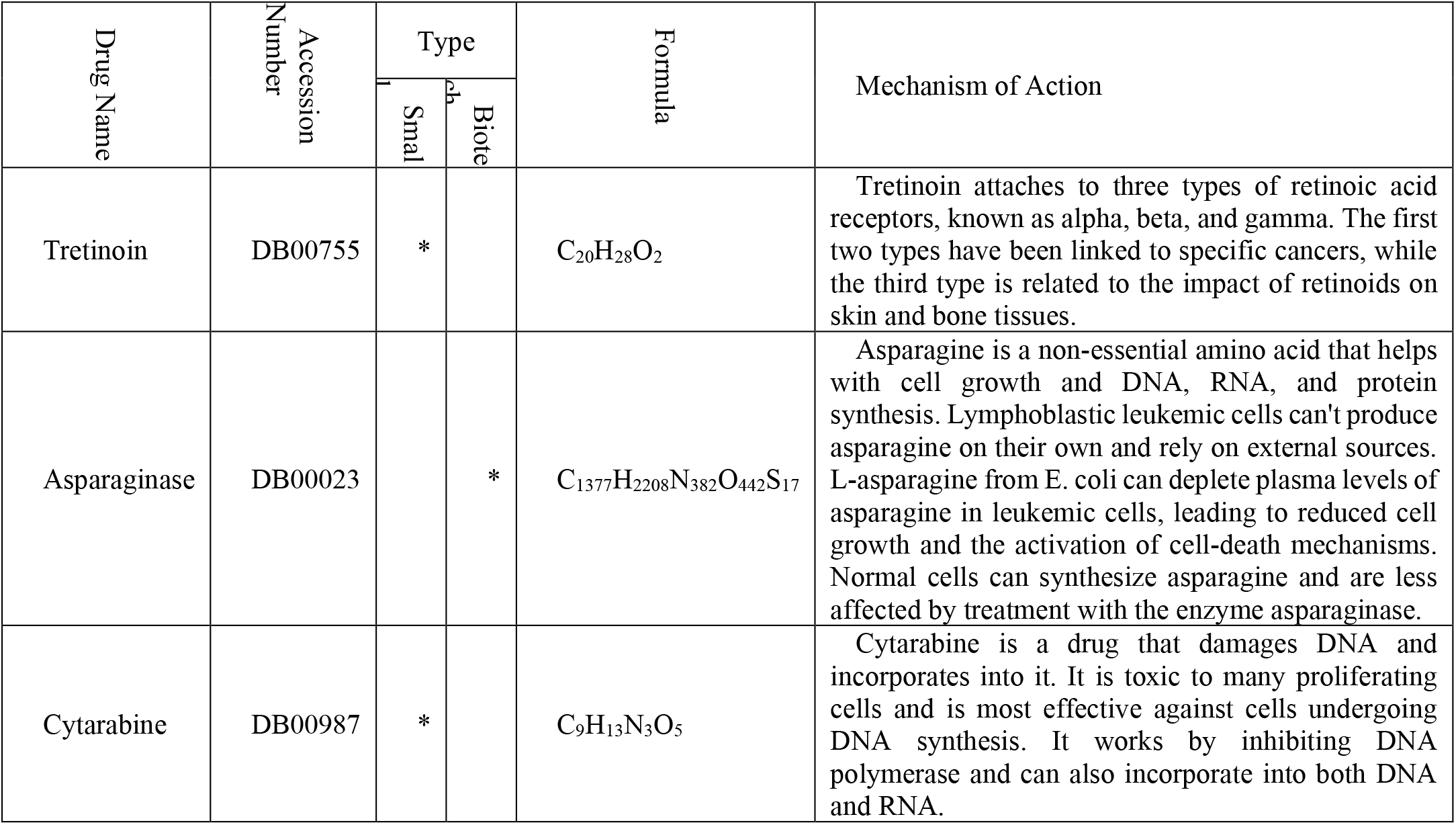
properties of proposed drugs as effective drugs to Leukemia management.

**Table 4:**
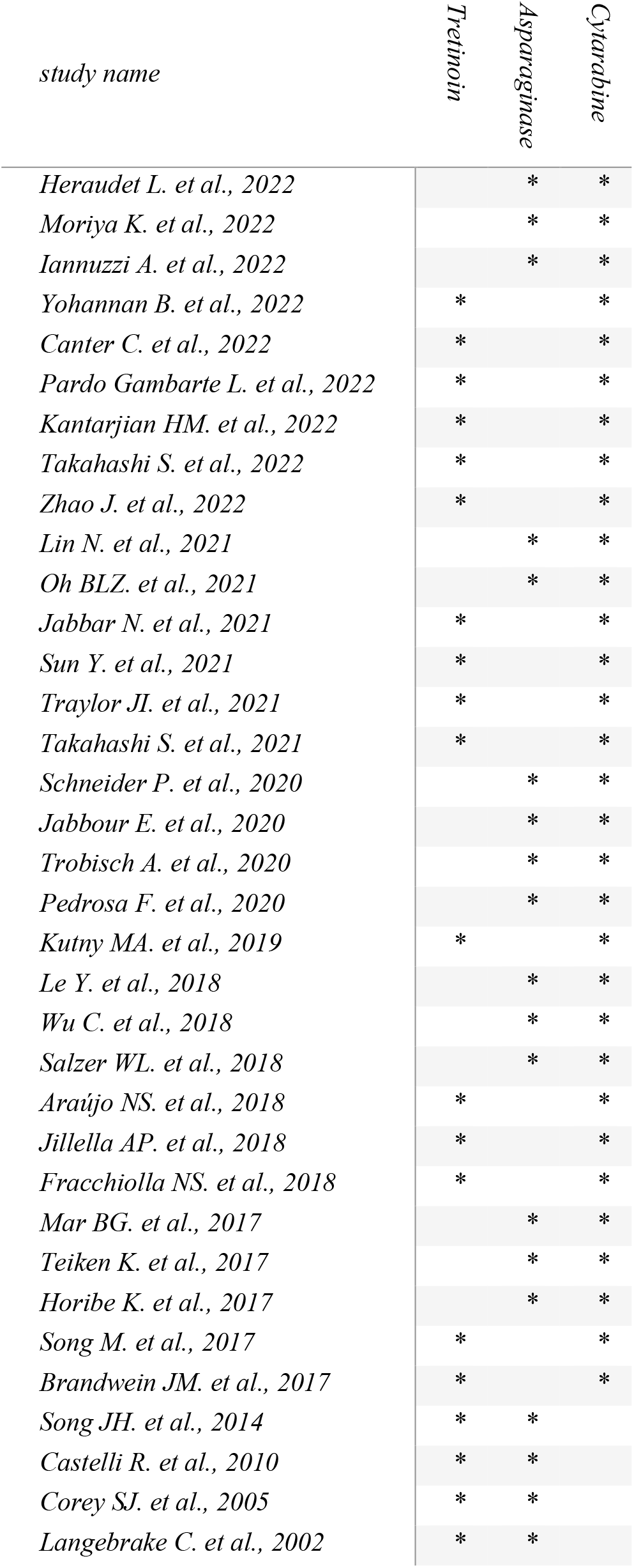
some important research studies for proposed drugs in Leukemia managements.

**Figure 4:**
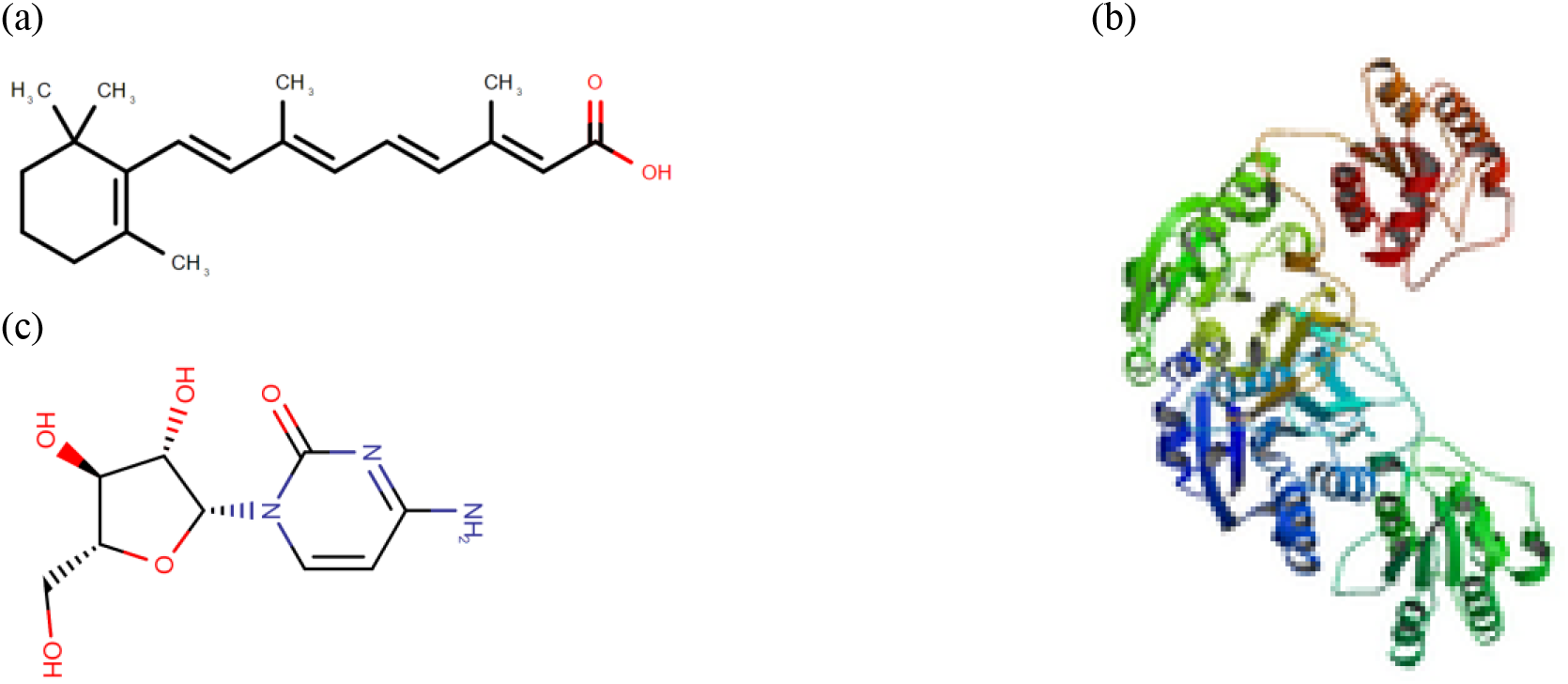
Drug structure for (a) Tretinoin, (b) Asparaginase, (c) Cytarabine from https://www.drugbank.com/.

### Stage III: Network Meta-Analysis

Figure 5 exhibits the p-values for human proteins/genes affected by Leukemia, whereas Figure 6 presents the p-values following the implementation of the third scenario. The efficacy of drugs chosen by the drug selection algorithm is demonstrated in Figure 7 through a radar chart, indicating the p-values between Leukemia and human proteins/genes after the administration of the selected drugs. Figure 8 illustrates the p-values between associations and targets using different interface features. P-values lower than .01 and .05 are depicted in green and blue, respectively. Each colored line represents the effectiveness of the corresponding drug in the given scenario.

**Figure 5:**
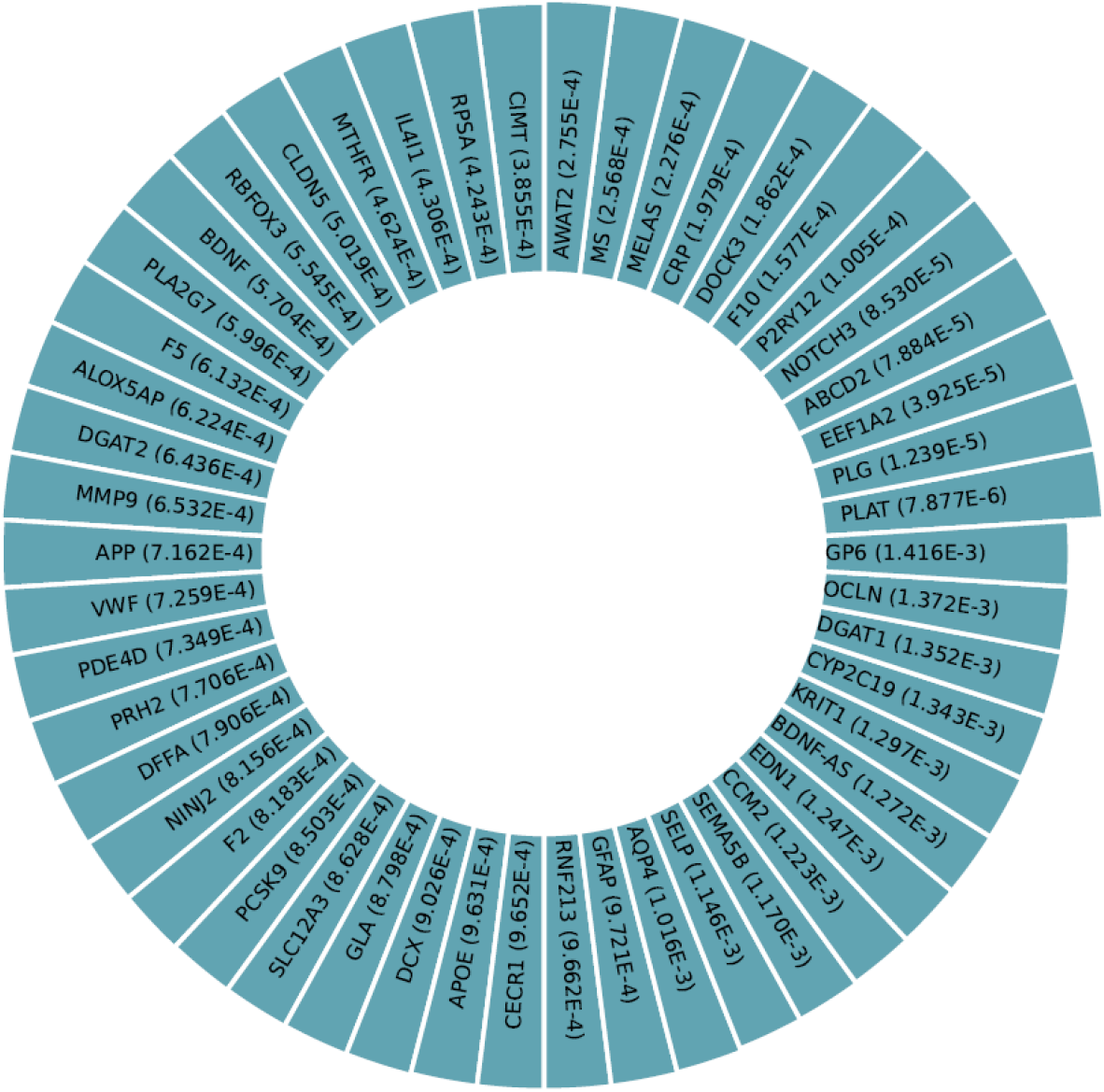
p-values between affected human proteins/genes and Leukemia.

**Figure 6:**
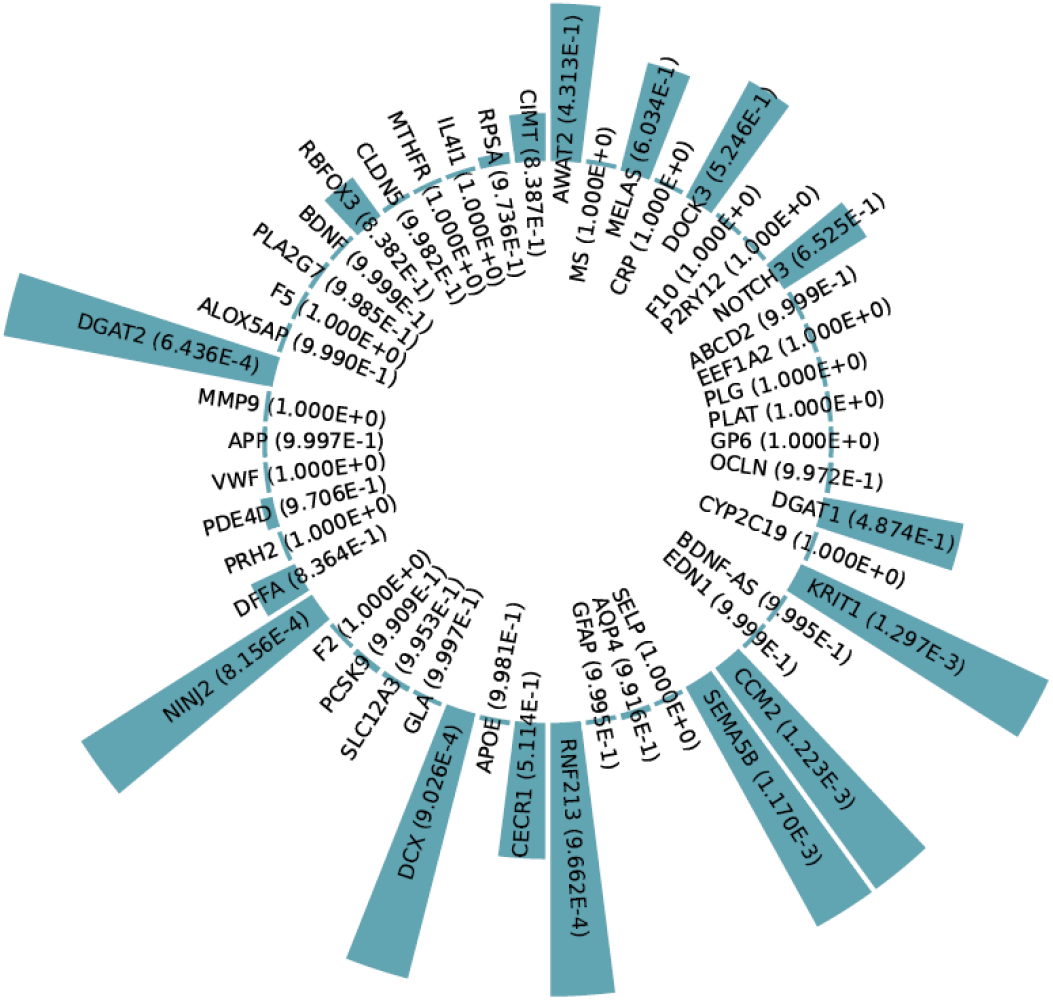
p-values between affected human proteins/genes and Leukemia after implementing Scenario 4.

**Figure 7:**
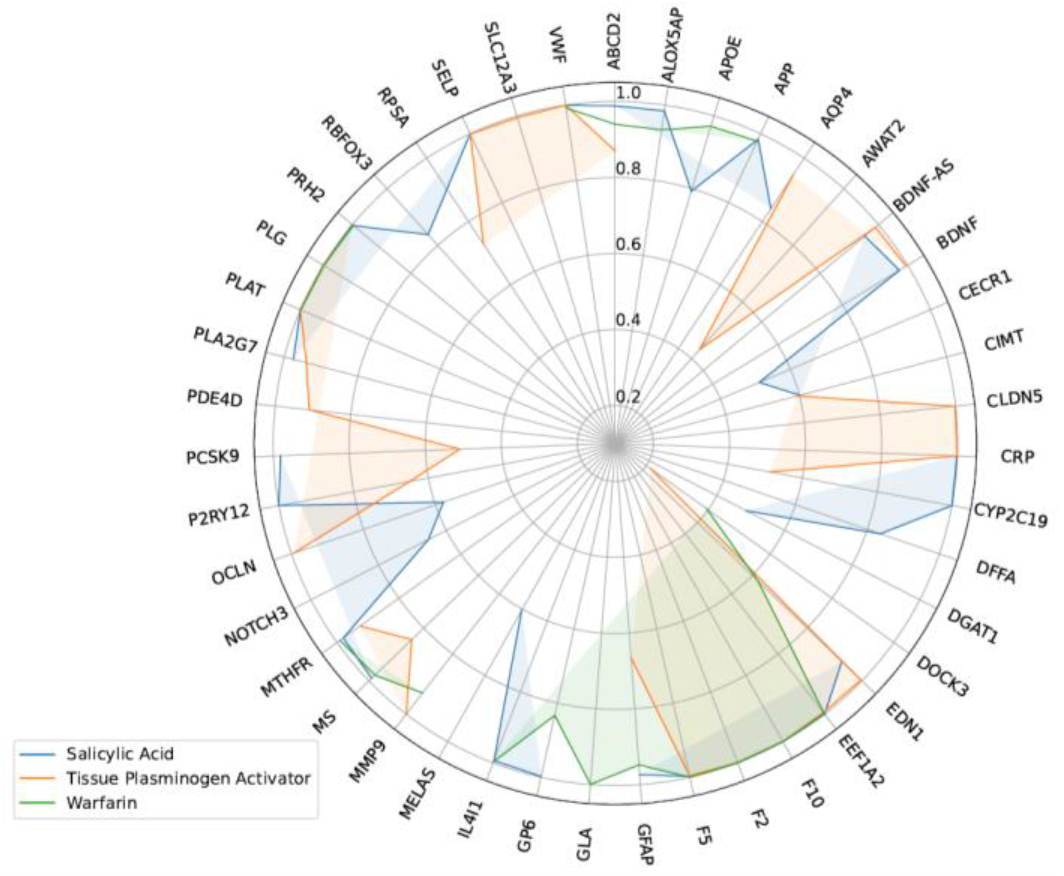
radar chart for p-values between Leukemia and affected proteins/genes, after consumption of each drug.

**Figure 8:**
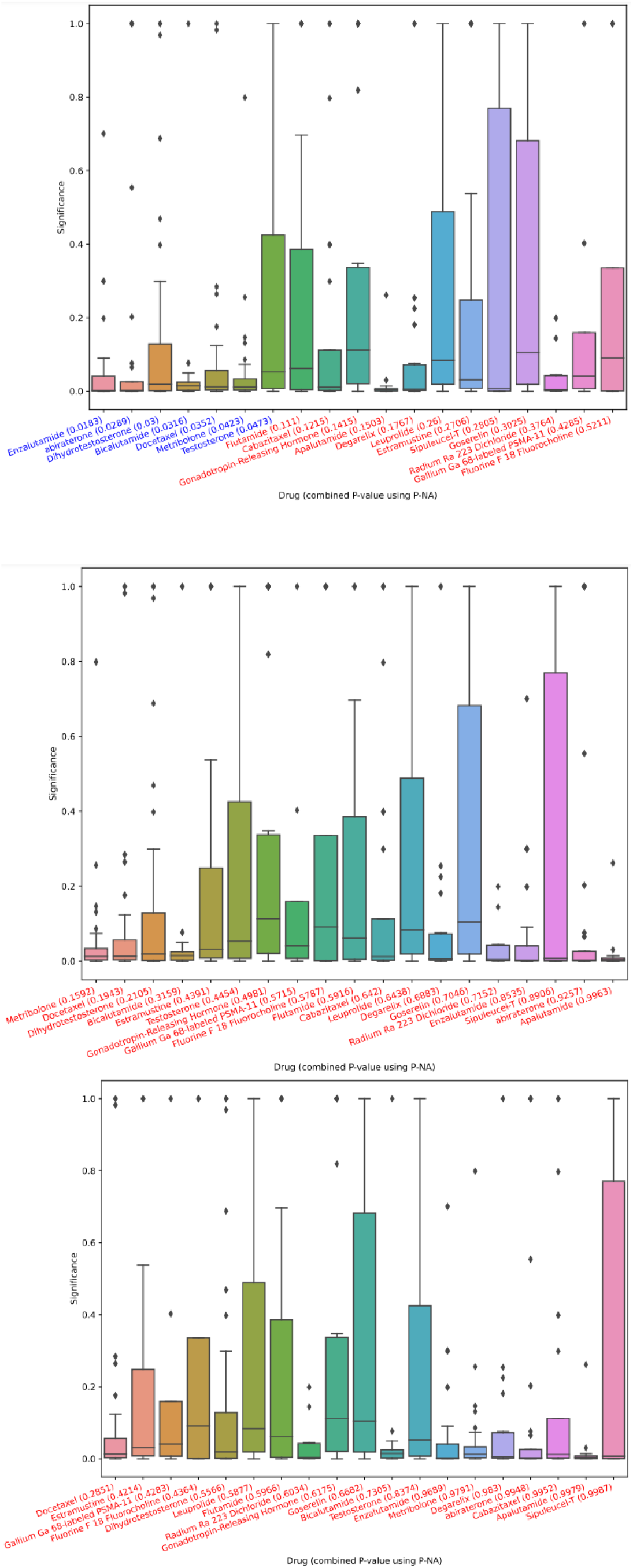
p-values between associations and target, using different interface features. (a) Overall, (b) after first drug from (a) is used, (c) after first drugs of (a) and (b) are used.

## DISCUSSION

Prescription drug information plays a crucial role in investigating the potential interactions, adverse effects, and risks associated with medications. To discern such details, various trustworthy online platforms like Medscape, WebMD, Drugs, and Drugbank were consulted to facilitate a comprehensive comparison of different drugs. Through these reliable databases, a thorough analysis of drug combinations was conducted, ultimately revealing the existence of certain interactions among specific medication pairs. However, it is important to note that the investigated websites have not identified any substantial interactions between Tretinoin, Asparaginase, and Cytarabine at present.

## CONCLUSION

Leukemia is a form of cancer that impacts the body’s blood-forming tissues, such as the bone marrow and lymphatic system. Typically, it affects the white blood cells, which serve as powerful infection fighters and normally grow and divide in an organized manner as needed by the body. By utilizing a combination of drugs, the treatment of leukemia can be enhanced. For acute leukemias, induction chemotherapy is often the initial step, followed by intensification or consolidation chemotherapy. Through the implementation of the RAIN protocol, our Graph Neural Network has recommended a combination of Tretinoin, Asparaginase, and Cytarabine to specifically target human genes and proteins associated with leukemia. Findings indicate a notable advancement in the treatment of leukemia.

## Data Availability

All data produced in the present study are available upon reasonable request to the authors

## Abbreviations

STROBE: Strengthening the Reporting of Observational studies in Epidemiology
PRISMA: Preferred Reporting Items for Systematic Reviews and Meta-Analysis
RAIN: Systematic Review and Artificial Intelligence Network Meta-Analysis

## Acknowledgements

By researchers in Bioinformatics, Computational Biology, Artificial Intelligence and Medicine.

## Authors’ contributions

HM contributed to design, AAK and MB native RL algorithm, HM, AAK, MB bio-statistical analysis; MB participated in most of the study steps. AAK used RL in exploring drug combinations. All authors have read and approved the content of the manuscript

## Availability of data and materials

Datasets are available through the corresponding author upon reasonable request.

## Ethics approval and consent to participate

Not applicable.

## Consent for publication

Not applicable.

## References

[1] “A PML/RARα direct target atlas redefines transcriptional deregulation in acute promyelocytic leukemia | Blood | American Society of Hematology.” https://ashpublications.org/blood/article/137/11/1503/463523/A-PML-RAR-direct-target-atlas-redefines (accessed Jul. 03, 2023).

[2] N. I. Noguera et al., “Acute Promyelocytic Leukemia: Update on the Mechanisms of Leukemogenesis, Resistance and on Innovative Treatment Strategies,” Cancers (Basel), vol. 11, no. 10, p. 1591, Oct. 2019, doi: 10.3390/cancers11101591.

[3] “PML-RARA–targeted DNA vaccine induces protective immunity in a mouse model of leukemia | Nature Medicine.” https://www.nature.com/articles/nm949 (accessed Jul. 03, 2023).

[4] S. Viswanadhapalli, K. V. Dileep, K. Y. J. Zhang, H. B. Nair, and R. K. Vadlamudi, “Targeting LIF/LIFR signaling in cancer,” Genes Dis, vol. 9, no. 4, pp. 973–980, Apr. 2021, doi: 10.1016/j.gendis.2021.04.003.

[5] E. Wrona, P. Potemski, F. Sclafani, and M. Borowiec, “Leukemia Inhibitory Factor: A Potential Biomarker and Therapeutic Target in Pancreatic Cancer,” Arch Immunol Ther Exp (Warsz), vol. 69, no. 1, p. 2, 2021, doi: 10.1007/s00005-021-00605-w.

[6] W. Tang et al., “LIF/LIFR oncogenic signaling is a novel therapeutic target in endometrial cancer,” Cell Death Discov., vol. 7, no. 1, Art. no. 1, Aug. 2021, doi: 10.1038/s41420-021-00603-z.

[7] C. Meyer et al., “The KMT2A recombinome of acute leukemias in 2023,” Leukemia, vol. 37, no. 5, Art. no. 5, May 2023, doi: 10.1038/s41375-023-01877-1.

[8] M. Górecki, I. Koziol, A. Kopystecka, J. Budzyńska, J. Zawitkowska, and M. Lejman, “Updates in KMT2A Gene Rearrangement in Pediatric Acute Lymphoblastic Leukemia,” Biomedicines, vol. 11, no. 3, Art. no. 3, Mar. 2023, doi: 10.3390/biomedicines11030821.

[9] G. C. Issa et al., “Predictors of outcomes in adults with acute myeloid leukemia and KMT2A rearrangements,” Blood Cancer J., vol. 11, no. 9, Art. no. 9, Sep. 2021, doi: 10.1038/s41408-021-00557-6.

[10] S. Goyama, G. Huang, M. Kurokawa, and J. C. Mulloy, “Posttranslational modifications of RUNX1 as potential anticancer targets,” Oncogene, vol. 34, no. 27, Art. no. 27, Jul. 2015, doi: 10.1038/onc.2014.305.

[11] J. Michaud et al., “Integrative analysis of RUNX1 downstream pathways and target genes,” BMC Genomics, vol. 9, no. 1, p. 363, Jul. 2008, doi: 10.1186/1471-2164-9-363.

[12] A. J. Ambinder and M. Levis, “Potential targeting of FLT3 acute myeloid leukemia,” Haematologica, vol. 106, no. 3, pp. 671–681, Jul. 2020, doi: 10.3324/haematol.2019.240754.

[13] N. Daver, S. Venugopal, and F. Ravandi, “FLT3 mutated acute myeloid leukemia: 2021 treatment algorithm,” Blood Cancer J., vol. 11, no. 5, Art. no. 5, May 2021, doi: 10.1038/s41408-021-00495-3.

[14] Y. Dasgupta et al., “Normal ABL1 is a tumor suppressor and therapeutic target in human and mouse leukemias expressing oncogenic ABL1 kinases,” Blood, vol. 127, no. 17, pp. 2131–2143, Apr. 2016, doi: 10.1182/blood-2015-11-681171.

[15] H. Lee, I. N. Basso, and D. D. H. Kim, “Target spectrum of the BCR-ABL tyrosine kinase inhibitors in chronic myeloid leukemia,” Int J Hematol, vol. 113, no. 5, pp. 632–641, May 2021, doi: 10.1007/s12185-021-03126-6.

[16] H. Zhao and M. W. Deininger, “Declaration of Bcr-Abl1 independence,” Leukemia, vol. 34, no. 11, Art. no. 11, Nov. 2020, doi: 10.1038/s41375-020-01037-9.

[17] “Targeted Therapies for Chronic Myeloid Leukemia.” https://www.cancer.org/cancer/types/chronic-myeloid-leukemia/treating/targeted-therapies.html (accessed Jul. 03, 2023).

[18] L. Mazzera et al., “MEK1/2 regulate normal BCR and ABL1 tumor-suppressor functions to dictate ATO response in TKI-resistant Ph+ leukemia,” Leukemia, pp. 1–15, Jun. 2023, doi: 10.1038/s41375-023-01940-x.

[19] H. Mu, X. Zhu, H. Jia, L. Zhou, and H. Liu, “Combination Therapies in Chronic Myeloid Leukemia for Potential Treatment-Free Remission: Focus on Leukemia Stem Cells and Immune Modulation,” Frontiers in Oncology, vol. 11, 2021, Accessed: Jul. 03, 2023. [Online]. Available: https://www.frontiersin.org/articles/10.3389/fonc.2021.643382

[20] K. Fousek et al., “CAR T-cells that target acute B-lineage leukemia irrespective of CD19 expression,” Leukemia, vol. 35, no. 1, Art. no. 1, Jan. 2021, doi: 10.1038/s41375-020-0792-2.

[21] S. Haque and S. R. Vaiselbuh, “CD19 Chimeric Antigen Receptor-Exosome Targets CD19 Positive B-lineage Acute Lymphocytic Leukemia and Induces Cytotoxicity,” Cancers (Basel), vol. 13, no. 6, p. 1401, Mar. 2021, doi: 10.3390/cancers13061401.

[22] A. Viardot and E. Sala, “Investigational immunotherapy targeting CD19 for the treatment of acute lymphoblastic leukemia,” Expert Opin Investig Drugs, vol. 30, no. 7, pp. 773–784, Jul. 2021, doi: 10.1080/13543784.2021.1928074.

[23] S.-K. Heo et al., “CD45dimCD34+CD38-CD133+ cells have the potential as leukemic stem cells in acute myeloid leukemia,” BMC Cancer, vol. 20, no. 1, p. 285, Apr. 2020, doi: 10.1186/s12885-020-06760-1.

[24] H. Herrmann et al., “Delineation of target expression profiles in CD34+/CD38- and CD34+/CD38+ stem and progenitor cells in AML and CML,” Blood Adv, vol. 4, no. 20, pp. 5118–5132, Oct. 2020, doi: 10.1182/bloodadvances.2020001742.

[25] K. Mattes et al., “CD34+ acute myeloid leukemia cells with low levels of reactive oxygen species show increased expression of stemness genes and can be targeted by the BCL2 inhibitor venetoclax,” Haematologica, vol. 105, no. 8, pp. e399–e403, Aug. 2020, doi: 10.3324/haematol.2019.229997.

[26] K. Liu and C. A. Tirado, “MECOM: A Very Interesting Gene Involved also in Lymphoid Malignancies,” J Assoc Genet Technol, vol. 45, no. 3, pp. 109–114, 2019.

[27] H. Kantarjian et al., “Acute myeloid leukemia: current progress and future directions,” Blood Cancer J, vol. 11, no. 2, p. 41, Feb. 2021, doi: 10.1038/s41408-021-00425-3.

[28] S. Kiehlmeier et al., “Identification of therapeutic targets of the hijacked super-enhancer complex in EVI1-rearranged leukemia,” Leukemia, vol. 35, no. 11, Art. no. 11, Nov. 2021, doi: 10.1038/s41375-021-01235-z.

[29] K.-H. T. Dao et al., “Efficacy of Ruxolitinib in Patients With Chronic Neutrophilic Leukemia and Atypical Chronic Myeloid Leukemia,” JCO, vol. 38, no. 10, pp. 1006–1018, Apr. 2020, doi: 10.1200/JCO.19.00895.

[30] J. E. Maxson et al., “Oncogenic CSF3R Mutations in Chronic Neutrophilic Leukemia and Atypical CML,” New England Journal of Medicine, vol. 368, no. 19, pp. 1781– 1790, May 2013, doi: 10.1056/NEJMoa1214514.

[31] E. Fabiani et al., “Mutational profile of ZBTB16-RARA-positive acute myeloid leukemia,” Cancer Med, vol. 10, no. 12, pp. 3839–3847, Jun. 2021, doi: 10.1002/cam4.3904.

[32] M. E. Matyskiela et al., “Cereblon Modulators Target ZBTB16 and Its Oncogenic Fusion Partners for Degradation via Distinct Structural Degrons,” ACS Chem Biol, vol. 15, no. 12, pp. 3149–3158, Dec. 2020, doi: 10.1021/acschembio.0c00674.

[33] Q. Wang et al., “Hyperthermia promotes degradation of the acute promyelocytic leukemia driver oncoprotein ZBTB16/RARα,” Acta Pharmacol Sin, vol. 44, no. 4, Art. no. 4, Apr. 2023, doi: 10.1038/s41401-022-01001-6.

[34] P. Xiang et al., “Elucidating the importance and regulation of key enhancers for human MEIS1 expression,” Leukemia, vol. 36, no. 8, Art. no. 8, Aug. 2022, doi: 10.1038/s41375-022-01602-4.

[35] M. Yao, Y. Gu, Z. Yang, K. Zhong, and Z. Chen, “MEIS1 and its potential as a cancer therapeutic target (Review),” Int J Mol Med, vol. 48, no. 3, p. 181, Sep. 2021, doi: 10.3892/ijmm.2021.5014.

[36] C. T. Collins and J. L. Hess, “Role of HOXA9 in leukemia: dysregulation, cofactors and essential targets,” Oncogene, vol. 35, no. 9, pp. 1090–1098, Mar. 2016, doi: 10.1038/onc.2015.174.

[37] G. F. Perini, G. N. Ribeiro, J. V. Pinto Neto, L. T. Campos, and N. Hamerschlak, “BCL-2 as therapeutic target for hematological malignancies,” Journal of Hematology & Oncology, vol. 11, no. 1, p. 65, May 2018, doi: 10.1186/s13045-018-0608-2.

[38] Y. Wei et al., “Targeting Bcl-2 Proteins in Acute Myeloid Leukemia,” Front Oncol, vol. 10, p. 584974, Nov. 2020, doi: 10.3389/fonc.2020.584974.

[39] “BCL2 Inhibitors Selectively Target Leukemia Stem Cells,” Cancer Network, Feb. 23, 2013. https://www.cancernetwork.com/view/bcl2-inhibitors-selectively-target-leukemia-stem-cells (accessed Jul. 03, 2023).

[40] Y. Yi and S. Ge, “Targeting the histone H3 lysine 79 methyltransferase DOT1L in MLL-rearranged leukemias,” Journal of Hematology & Oncology, vol. 15, no. 1, p. 35, Mar. 2022, doi: 10.1186/s13045-022-01251-1.

[41] E. R. Barry, G. N. Corry, and T. P. Rasmussen, “Targeting DOT1L action and interactions in leukemia: the role of DOT1L in transformation and development,” Expert Opin Ther Targets, vol. 14, no. 4, pp. 405–418, Apr. 2010, doi: 10.1517/14728221003623241.

[42] C. M. McLean, I. D. Karemaker, and F. van Leeuwen, “The emerging roles of DOT1L in leukemia and normal development,” Leukemia, vol. 28, no. 11, Art. no. 11, Nov. 2014, doi: 10.1038/leu.2014.169.

[43] B. Zhou et al., “WT1 facilitates the self-renewal of leukemia-initiating cells through the upregulation of BCL2L2: WT1-BCL2L2 axis as a new acute myeloid leukemia therapy target,” Journal of Translational Medicine, vol. 18, no. 1, p. 254, Jun. 2020, doi: 10.1186/s12967-020-02384-y.

[44] A. G. Chapuis et al., “T cell receptor gene therapy targeting WT1 prevents acute myeloid leukemia relapse post-transplant,” Nat Med, vol. 25, no. 7, Art. no. 7, Jul. 2019, doi: 10.1038/s41591-019-0472-9.

[45] S. Piya et al., “Targeting the NOTCH1-MYC-CD44 axis in leukemia-initiating cells in T-ALL,” Leukemia, vol. 36, no. 5, Art. no. 5, May 2022, doi: 10.1038/s41375-022-01516-1.

[46] C. Wang et al., “Alternative approaches to target Myc for cancer treatment,” Sig Transduct Target Ther, vol. 6, no. 1, Art. no. 1, Mar. 2021, doi: 10.1038/s41392-021-00500-y.

[47] O. de Barrios, A. Meler, and M. Parra, “MYC’s Fine Line Between B Cell Development and Malignancy,” Cells, vol. 9, no. 2, p. 523, Feb. 2020, doi: 10.3390/cells9020523.

[48] B. A. Vishwakarma, K. O. Gudmundsson, K. Oakley, Y. Han, and Y. Du, “Insertional mutagenesis identifies cooperation between Setbp1 and Mllt3 in inducing myeloid leukemia development,” Leukemia, vol. 33, no. 8, Art. no. 8, Aug. 2019, doi: 10.1038/s41375-019-0445-5.

[49] C. R. Schmidt et al., “BCOR Binding to MLL-AF9 Is Essential for Leukemia via Altered EYA1, SIX, and MYC Activity,” Blood Cancer Discov, vol. 1, no. 2, pp. 162– 177, Sep. 2020, doi: 10.1158/2643-3230.BCD-20-0036.

[50] F. El Chaer, M. Keng, and K. K. Ballen, “MLL-Rearranged Acute Lymphoblastic Leukemia,” Curr Hematol Malig Rep, vol. 15, no. 2, pp. 83–89, Apr. 2020, doi: 10.1007/s11899-020-00582-5.

[51] R. Ranieri et al., “Current status and future perspectives in targeted therapy of NPM1-mutated AML,” Leukemia, vol. 36, no. 10, Art. no. 10, Oct. 2022, doi: 10.1038/s41375-022-01666-2.

[52] Y. Shi, Y. Xue, C. Wang, and L. Yu, “Nucleophosmin 1: from its pathogenic role to a tantalizing therapeutic target in acute myeloid leukemia,” Hematology, vol. 27, no. 1, pp. 609–619, Dec. 2022, doi: 10.1080/16078454.2022.2067939.

[53] M. O. Forgione, B. J. McClure, D. T. Yeung, L. N. Eadie, and D. L. White, “MLLT10 rearranged acute leukemia: Incidence, prognosis, and possible therapeutic strategies,” Genes Chromosomes Cancer, Jul. 2020, doi: 10.1002/gcc.22887.

[54] T. Mahmoudi et al., “The Leukemia-Associated Mllt10/Af10-Dot1l Are Tcf4/β-Catenin Coactivators Essential for Intestinal Homeostasis,” PLOS Biology, vol. 8, no. 11, p. e1000539, Nov. 2010, doi: 10.1371/journal.pbio.1000539.

[55] E. R. Vagapova, P. V. Spirin, T. D. Lebedev, and V. S. Prassolov, “The Role of TAL1 in Hematopoiesis and Leukemogenesis,” Acta Naturae, vol. 10, no. 1, pp. 15–23, 2018.

[56] C. Smith et al., “TAL1 activation in T-cell acute lymphoblastic leukemia: a novel oncogenic 3’ neo-enhancer,” Haematologica, vol. 108, no. 5, pp. 1259– 1271, May 2023, doi: 10.3324/haematol.2022.281583.

[57] K. Karjalainen et al., “Targeting IL11 receptor in leukemia and lymphoma: A functional ligand-directed study and hematopathology analysis of patient-derived specimens,” Clinical Cancer Research, vol. 21, no. 13, pp. 3041–3051, Jul. 2015, doi: 10.1158/1078-0432.CCR-13-3059.

[58] A. Charlet et al., “The IL-3, IL-5, and GM-CSF common receptor beta chain mediates oncogenic activity of FLT3-ITD-positive AML,” Leukemia, vol. 36, no. 3, Art. no. 3, Mar. 2022, doi: 10.1038/s41375-021-01462-4.

[59] V. Pierini et al., “A Novel t(5;7)(q31;q21)/CDK6::IL3 in Immature T-cell Acute Lymphoblastic Leukemia With IL3 Expression and Eosinophilia,” Hemasphere, vol. 6, no. 11, p. e795, Oct. 2022, doi: 10.1097/HS9.0000000000000795.

[60] Y. Tan et al., “A PML/RARα direct target atlas redefines transcriptional deregulation in acute promyelocytic leukemia,” Blood, The Journal of the American Society of Hematology, vol. 137, no. 11, pp. 1503–1516, 2021.

[61] C. EJ. Downes et al., “JAK2 Alterations in Acute Lymphoblastic Leukemia: Molecular Insights for Superior Precision Medicine Strategies,” Frontiers in Cell and Developmental Biology, vol. 10, 2022, Accessed: Jul. 03, 2023. [Online]. Available: https://www.frontiersin.org/articles/10.3389/fcell.2022.942053

[62] N. Gangat, N. Szuber, A. Pardanani, and A. Tefferi, “JAK2 unmutated erythrocytosis: current diagnostic approach and therapeutic views,” Leukemia, vol. 35, no. 8, Art. no. 8, Aug. 2021, doi: 10.1038/s41375-021-01290-6.

[63] E. S. Fasouli and E. Katsantoni, “JAK-STAT in Early Hematopoiesis and Leukemia,” Front Cell Dev Biol, vol. 9, p. 669363, May 2021, doi: 10.3389/fcell.2021.669363.

[64] D. Hoelzer, “Anti-CD22 therapy in acute lymphoblastic leukaemia,” The Lancet Oncology, vol. 13, no. 4, pp. 329– 331, Apr. 2012, doi: 10.1016/S1470-2045(12)70010-4.

[65] N. N. Shah et al., “Characterization of CD22 expression in acute lymphoblastic leukemia,” Pediatr Blood Cancer, vol. 62, no. 6, pp. 964–969, Jun. 2015, doi: 10.1002/pbc.25410.

[66] S. Al-Harbi, M. Aljurf, M. Mohty, F. Almohareb, and S. O. A. Ahmed, “An update on the molecular pathogenesis and potential therapeutic targeting of AML with t(8;21)(q22;q22.1);RUNX1-RUNX1T1,” Blood Adv, vol. 4, no. 1, pp. 229–238, Jan. 2020, doi: 10.1182/bloodadvances.2019000168.

[67] N. H. E. Darwish et al., “Acute myeloid leukemia stem cell markers in prognosis and targeted therapy: potential impact of BMI-1, TIM-3 and CLL-1,” Oncotarget, vol. 7, no. 36, pp. 57811–57820, Sep. 2016, doi: 10.18632/oncotarget.11063.

[68] S. A. Mariani, V. Minieri, M. De Dominici, I. Iacobucci, L. F. Peterson, and B. Calabretta, “CDKN2A-independent role of BMI1 in promoting growth and survival of Ph+ acute lymphoblastic leukemia,” Leukemia, vol. 30, no. 8, Art. no. 8, Aug. 2016, doi: 10.1038/leu.2016.70.

[69] L. Rouhigharabaei et al., “BMI1, the polycomb-group gene, is recurrently targeted by genomic rearrangements in progressive B-cell leukemia/lymphoma,” Genes Chromosomes Cancer, vol. 52, no. 10, pp. 928–944, Oct. 2013, doi: 10.1002/gcc.22088.

[70] B. S. Chhikara and K. Parang, “Development of cytarabine prodrugs and delivery systems for leukemia treatment,” Expert opinion on drug delivery, vol. 7, no. 12, pp. 1399– 1414, 2010.

[71] B. Löwenberg et al., “Cytarabine Dose for Acute Myeloid Leukemia,” New England Journal of Medicine, vol. 364, no. 11, pp. 1027–1036, Mar. 2011, doi: 10.1056/NEJMoa1010222.

[72] N. Daver, A. H. Wei, D. A. Pollyea, A. T. Fathi, P. Vyas, and C. D. DiNardo, “New directions for emerging therapies in acute myeloid leukemia: the next chapter,” Blood Cancer J., vol. 10, no. 10, Art. no. 10, Oct. 2020, doi: 10.1038/s41408-020-00376-1.

[73] H. Wang et al., “Venetoclax plus 3 + 7 daunorubicin and cytarabine chemotherapy as first-line treatment for adults with acute myeloid leukaemia: a multicentre, single-arm, phase 2 trial,” The Lancet Haematology, vol. 9, no. 6, pp. e415–e424, Jun. 2022, doi: 10.1016/S2352-3026(22)00106-5.

[74] H. A. Blair, “Daunorubicin/Cytarabine Liposome: A Review in Acute Myeloid Leukaemia,” Drugs, vol. 78, no. 18, pp. 1903–1910, 2018, doi: 10.1007/s40265-018-1022-3.

[75] Q. Gong, L. Zhou, S. Xu, X. Li, Y. Zou, and J. Chen, “High Doses of Daunorubicin during Induction Therapy of Newly Diagnosed Acute Myeloid Leukemia: A Systematic Review and Meta-Analysis of Prospective Clinical Trials,” PLoS One, vol. 10, no. 5, p. e0125612. May 2015, doi: 10.1371/journal.pone.0125612.

[76] R. Kamojjala and B. Bostrom, “Allopurinol to Prevent Mercaptopurine Adverse Effects in Children and Young Adults With Acute Lymphoblastic Leukemia,” Journal of Pediatric Hematology/Oncology, vol. 43, no. 3, p. 95, Apr. 2021, doi: 10.1097/MPH.0000000000002117.

[77] A. Østergaard et al., “Acute lymphoblastic leukemia and down syndrome: 6-mercaptopurine and methotrexate metabolites during maintenance therapy,” Leukemia, vol. 35, no. 3, Art. no. 3, Mar. 2021, doi: 10.1038/s41375-020-0946-2.s

[78] J. H. Burchenal et al., “Clinical evaluation of a new antimetabolite, 6-mercaptopurine, in the treatment of leukemia and allied diseases,” Blood, vol. 8, no. 11, pp. 965–999, 1953.

[79] W. Yang et al., “Pulse therapy with vincristine and dexamethasone for childhood acute lymphoblastic leukaemia (CCCG-ALL-2015): an open-label, multicentre, randomised, phase 3, non-inferiority trial,” The Lancet Oncology, vol. 22, no. 9, pp. 1322–1332, Sep. 2021, doi: 10.1016/S1470-2045(21)00328-4.

[80] J. Škubník, V. S. Pavlíčková, T. Ruml, and S. Rimpelová, “Vincristine in Combination Therapy of Cancer: Emerging Trends in Clinics,” Biology (Basel), vol. 10, no. 9, p. 849, Aug. 2021, doi: 10.3390/biology10090849.

[81] H. T. Masoumi, M. Hadjibabaie, M. Zarif-Yeganeh, B. Khajeh, and A. Ghavamzadeh, “Treatment of vincristine-induced ileus with metoclopramide: A case report,” J Oncol Pharm Pract, vol. 25, no. 2, pp. 507–511, Mar. 2019, doi: 10.1177/1078155217746228.

[82] M.-W. Chao et al., “The synergic effect of vincristine and vorinostat in leukemia in vitro and in vivo,” J Hematol Oncol, vol. 8, p. 82, Jul. 2015, doi: 10.1186/s13045-015-0176-7.

[83] R. Gorlick, E. Goker, T. Trippett, M. Waltham, D. Banerjee, and J. R. Bertino, “Intrinsic and Acquired Resistance to Methotrexate in Acute Leukemia,” N Engl J Med, vol. 335, no. 14, pp. 1041–1048, Oct. 1996, doi: 10.1056/NEJM199610033351408.

[84] E. Lopez-Lopez and W. E. Evans, “New insights into methotrexate accumulation in leukemia cells in vivo,” Mol Cell Oncol, vol. 8, no. 1, p. 1865086, doi: 10.1080/23723556.2020.1865086.

[85] V. J. Forster, A. McDonnell, R. Theobald, and J. A. McKay, “Effect of methotrexate/vitamin B12 on DNA methylation as a potential factor in leukemia treatment-related neurotoxicity,” Epigenomics, vol. 9, no. 9, pp. 1205–1218, Sep. 2017, doi: 10.2217/epi-2016-0165.

[86] L. N. Toksvang, S. H. R. Lee, J. J. Yang, and K. Schmiegelow, “Maintenance therapy for acute lymphoblastic leukemia: basic science and clinical translations,” Leukemia, vol. 36, no. 7, Art. no. 7, Jul. 2022, doi: 10.1038/s41375-022-01591-4.

[87] T. Sakura et al., “High-dose methotrexate therapy significantly improved survival of adult acute lymphoblastic leukemia: a phase III study by JALSG,” Leukemia, vol. 32, no. 3, Art. no. 3, Mar. 2018, doi: 10.1038/leu.2017.283.

[88] D. W. Beelen et al., “Treosulfan or busulfan plus fludarabine as conditioning treatment before allogeneic haemopoietic stem cell transplantation for older patients with acute myeloid leukaemia or myelodysplastic syndrome (MC-FludT.14/L): a randomised, noninferiority, phase 3 trial,” The Lancet Haematology, vol. 7, no. 1, pp. e28–e39, Jan. 2020, doi: 10.1016/S2352-3026(19)30157-7.

[89] K. Begna, A. Abdelatif, S. Schwager, C. Hanson, A. Pardanani, and A. Tefferi, “Busulfan for the treatment of myeloproliferative neoplasms: the Mayo Clinic experience,” Blood Cancer Journal, vol. 6, no. 5, Art. no. 5, May 2016, doi: 10.1038/bcj.2016.34.

[90] R. Patel and P. Tadi, “Busulfan,” in StatPearls, Treasure Island (FL): StatPearls Publishing, 2023. Accessed: Jul. 03, 2023. [Online]. Available: http://www.ncbi.nlm.nih.gov/books/NBK555986/

[91] H. Zhang et al., “Busulfan Plus Cyclophosphamide Versus Total Body Irradiation Plus Cyclophosphamide for Adults Acute B Lymphoblastic Leukemia: An Open-Label, Multicenter, Phase III Trial,” JCO, vol. 41, no. 2, pp. 343– 353, Jan. 2023, doi: 10.1200/JCO.22.00767.

[92] P. J. Tutschka, E. A. Copelan, and J. P. Klein, “Bone Marrow Transplantation for Leukemia Following a New Busulfan and Cyclophosphamide Regimen,” Blood, vol. 70, no. 5, pp. 1382–1388, Nov. 1987, doi: 10.1182/blood.V70.5.1382.1382.

[93] A. Moignet et al., “Cyclophosphamide as a first-line therapy in LGL leukemia,” Leukemia, vol. 28, no. 5, pp. 1134–1136, May 2014, doi: 10.1038/leu.2013.359.

[94] Y. Zhao, H. Song, and L. Ni, “Cyclophosphamide for the treatment of acute lymphoblastic leukemia: A protocol for systematic review,” Medicine, vol. 98, no. 5, Feb. 2019, doi: 10.1097/MD.0000000000014293.

[95] H. A. Sherif, A. Magdy, H. A. Elshesheni, S. M. Ramadan, and R. A. Rashed, “Treatment outcome of doxorubicin versus idarubicin in adult acute myeloid leukemia,” Leuk Res Rep, vol. 16, p. 100272, Oct. 2021, doi: 10.1016/j.lrr.2021.100272.

[96] S. E. Lipshultz et al., “The Effect of Dexrazoxane on Myocardial Injury in Doxorubicin-Treated Children with Acute Lymphoblastic Leukemia,” New England Journal of Medicine, vol. 351, no. 2, pp. 145–153, Jul. 2004, doi: 10.1056/NEJMoa035153.

[97] E. Moles et al., “Delivery of PEGylated liposomal doxorubicin by bispecific antibodies improves treatment in models of high-risk childhood leukemia,” Science Translational Medicine, vol. 15, no. 696, p. eabm1262. May 2023, doi: 10.1126/scitranslmed.abm1262.

[98] K. R. Juluri, C. Siu, and R. D. Cassaday, “Asparaginase in the Treatment of Acute Lymphoblastic Leukemia in Adults: Current Evidence and Place in Therapy,” Blood Lymphat Cancer, vol. 12, pp. 55–79, May 2022, doi: 10.2147/BLCTT.S342052.

[99] S. Gupta et al., “Impact of Asparaginase Discontinuation on Outcome in Childhood Acute Lymphoblastic Leukemia: A Report From the Children’s Oncology Group,” JCO, vol. 38, no. 17, pp. 1897–1905, Jun. 2020, doi: 10.1200/JCO.19.03024.

[100] K. Hlozkova et al., “Metabolic profile of leukemia cells influences treatment efficacy of L-asparaginase,” BMC Cancer, vol. 20, p. 526, Jun. 2020, doi: 10.1186/s12885-020-07020-y.

[101] E. Damery, D. A. Solimando, and J. A. Waddell, “Arsenic Trioxide and Tretinoin (AsO/ATRA) for Acute Promyelocytic Leukemia (APL),” Hosp Pharm, vol. 51, no. 8, pp. 628–632, Sep. 2016, doi: 10.1310/hpj5108-628.

[102] F. Lo-Coco et al., “Retinoic acid and arsenic trioxide for acute promyelocytic leukemia,” New England Journal of Medicine, vol. 369, no. 2, pp. 111–121, 2013.

[103] R. P. Warrell et al., “Differentiation Therapy of Acute Promyelocytic Leukemia with Tretinoin (All-trans-Retinoic Acid),” New England Journal of Medicine, vol. 324, no. 20, pp. 1385–1393, May 1991, doi: 10.1056/NEJM199105163242002.

[104] N. Iqbal and N. Iqbal, “Imatinib: A Breakthrough of Targeted Therapy in Cancer,” Chemother Res Pract, vol. 2014, p. 357027, 2014, doi: 10.1155/2014/357027.

[105] A. Hochhaus et al., “Long-Term Outcomes of Imatinib Treatment for Chronic Myeloid Leukemia,” New England Journal of Medicine, vol. 376, no. 10, pp. 917–927, Mar. 2017, doi: 10.1056/NEJMoa1609324.

[106] H. Wang, X. Xiao, Q. Xiao, Y. Lu, and Y. Wu, “The efficacy and safety of daunorubicin versus idarubicin combined with cytarabine for induction therapy in acute myeloid leukemia,” Medicine (Baltimore), vol. 99, no. 24, p. e20094, Jun. 2020, 10.1097/MD.0000000000020094.

[107] T. M. Kadia et al., “Venetoclax plus intensive chemotherapy with cladribine, idarubicin, and cytarabine in patients with newly diagnosed acute myeloid leukaemia or high-risk myelodysplastic syndrome: a cohort from a single-centre, single-arm, phase 2 trial,” The Lancet Haematology, vol. 8, no. 8, pp. e552–e561, Aug. 2021, doi: 10.1016/S2352-3026(21)00192-7.

[108] G. H. Jackson et al., “The use of an all oral chemotherapy (idarubicin and etoposide) in the treatment of acute myeloid leukaemia in the elderly: a report of toxicity and efficacy,” Leukemia, vol. 11, no. 8, Art. no. 8, Aug. 1997, doi: 10.1038/sj.leu.2400726.

[109] A. Im et al., “Mitoxantrone and Etoposide for the Treatment of Acute Myeloid Leukemia Patients in First Relapse,” Oncol Res, vol. 24, no. 2, pp. 73–80, Jun. 2016, doi: 10.3727/096504016X14586627440156.

[110] W. Zhang, P. Gou, J.-M. Dupret, C. Chomienne, and F. Rodrigues-Lima, “Etoposide, an anticancer drug involved in therapy-related secondary leukemia: Enzymes at play,” Transl Oncol, vol. 14, no. 10, p. 101169, Jul. 2021, doi: 10.1016/j.tranon.2021.101169.

[111] S. Ezoe, “Secondary Leukemia Associated with the Anti-Cancer Agent, Etoposide, a Topoisomerase II Inhibitor,” Int J Environ Res Public Health, vol. 9, no. 7, pp. 2444– 2453, Jul. 2012, doi: 10.3390/ijerph9072444.

[112] L. N. Toksvang et al., “Thiopurine Enhanced ALL Maintenance (TEAM): study protocol for a randomized study to evaluate the improvement in disease-free survival by adding very low dose 6-thioguanine to 6-mercaptopurine/methotrexate-based maintenance therapy in pediatric and adult patients (0–45 years) with newly diagnosed B-cell precursor or T-cell acute lymphoblastic leukemia treated according to the intermediate risk-high group of the ALLTogether1 protocol,” BMC Cancer, vol. 22, 2022, doi: 10.1186/s12885-022-09522-3.

[113] M. Stanulla et al., “Hepatic sinusoidal obstruction syndrome and short-term application of 6-thioguanine in pediatric acute lymphoblastic leukemia,” Leukemia, vol. 35, no. 9, Art. no. 9, Sep. 2021, doi: 10.1038/s41375-021-01203-7.

[114] S. A. Mian, S. U. Khan, A. Hussain, A. Rauf, E. Ahmed, and J. Jang, “Molecular Modelling of Optical Biosensor Phosphorene-Thioguanine for Optimal Drug Delivery in Leukemia Treatment,” Cancers, vol. 14, no. 3, Art. no. 3, Jan. 2022, doi: 10.3390/cancers14030545.

[115] L. Chen et al., “Arsenic trioxide replacing or reducing chemotherapy in consolidation therapy for acute promyelocytic leukemia (APL2012 trial),” Proceedings of the National Academy of Sciences, vol. 118, no. 6, p. e2020382118, Feb. 2021, doi: 10.1073/pnas.2020382118.

[116] M. A. Kutny et al., “Assessment of Arsenic Trioxide and All-trans Retinoic Acid for the Treatment of Pediatric Acute Promyelocytic Leukemia,” JAMA Oncol, vol. 8, no. 1, pp. 1–9, Jan. 2022, doi: 10.1001/jamaoncol.2021.5206.

[117] A. Janssens, M. Boogaerts, and G. Verhoef, “Development of fludarabine formulations in the treatment of chronic lymphocytic leukemia,” Drug Des Devel Ther, vol. 3, pp. 241–252, Dec. 2009.

[118] R. Szász, B. Telek, and Á. Illés, “Fludarabine-Cyclophosphamide-Rituximab Treatment in Chronic Lymphocytic Leukemia, Focusing on Long Term Cytopenias Before and After the Era of Targeted Therapies,” Pathol Oncol Res, vol. 27, p. 1609742, Apr. 2021, doi: 10.3389/pore.2021.1609742.

[119] A. D. Bulgar et al., “Targeting base excision repair suggests a new therapeutic strategy of fludarabine for the treatment of chronic lymphocytic leukemia,” Leukemia, vol. 24, no. 10, Art. no. 10, Oct. 2010, doi: 10.1038/leu.2010.166.

[120] D. Kuron et al., “Amsacrine-based induction therapy in AML patients with cardiac comorbidities: a retrospective single-center analysis,” Ann Hematol, vol. 102, no. 4, pp. 755–760, Apr. 2023, doi: 10.1007/s00277-023-05111-x.

[121] K. Mezger et al., “Amsacrine combined with etoposide and methylprednisolone is a feasible and safe component in first-line intensified treatment of pediatric patients with high-risk acute lymphoblastic leukemia in CoALL08-09 trial,” Pediatr Blood Cancer, vol. 69, no. 12, p. e29997, Dec. 2022, doi: 10.1002/pbc.29997.

[122] H. Dombret and C. Gardin, “An update of current treatments for adult acute myeloid leukemia,” Blood, vol. 127, no. 1, pp. 53–61, Jan. 2016, doi: 10.1182/blood-2015-08-604520.

[123] Z. A. Arlin et al., “A new regimen of amsacrine with highdose cytarabine is safe and effective therapy for acute leukemia,” J Clin Oncol, vol. 5, no. 3, pp. 371–375, Mar. 1987, doi: 10.1200/JCO.1987.5.3.371.

[124] D. A. Vorobiof, G. Falkson, M. A. Coccia-Portugal, and A. P. S. Terblanche, “Mitoxantrone in the treatment of acute leukemia,” Invest New Drugs, vol. 5, no. 4, pp. 383–388, Dec. 1987, doi: 10.1007/BF00169980.

[125] “Mitoxantrone,” in LiverTox: Clinical and Research Information on Drug-Induced Liver Injury, Bethesda (MD): National Institute of Diabetes and Digestive and Kidney Diseases, 2012. Accessed: Jul. 03, 2023. [Online]. Available: http://www.ncbi.nlm.nih.gov/books/NBK547931/

[126] R. Di Francia et al., “Response and Toxicity to Cytarabine Therapy in Leukemia and Lymphoma: From Dose Puzzle to Pharmacogenomic Biomarkers,” Cancers (Basel), vol. 13, no. 5, p. 966, Feb. 2021, doi: 10.3390/cancers13050966.

[127] J. M. Rowe, “The ‘7+3’ regimen in acute myeloid leukemia,” Haematologica, vol. 107, no. 1, p. 3, Jan. 2022, doi: 10.3324/haematol.2021.280161.

[128] M. M. Patnaik, T. I. Mughal, C. Brooks, R. Lindsay, and N. Pemmaraju, “Targeting CD123 in hematologic malignancies: identifying suitable patients for targeted therapy,” Leukemia & Lymphoma, vol. 62, no. 11, pp. 2568–2586, Sep. 2021, doi: 10.1080/10428194.2021.1927021.

[129] A. Renneville, M. M. Patnaik, O. Chan, E. Padron, and E. Solary, “Increasing recognition and emerging therapies argue for dedicated clinical trials in chronic myelomonocytic leukemia,” Leukemia, vol. 35, no. 10, Art. no. 10, Oct. 2021, doi: 10.1038/s41375-021-01330-1.

[130] J. H. Black, J. A. McCubrey, M. C. Willingham, J. Ramage, D. E. Hogge, and A. E. Frankel, “Diphtheria toxin-interleukin-3 fusion protein (DT388IL3) prolongs disease-free survival of leukemic immunocompromised mice,” Leukemia, vol. 17, no. 1, Art. no. 1, Jan. 2003, doi: 10.1038/sj.leu.2402744.

[131] “Thrombocytopenia in Leukemia,” Medscape. http://www.medscape.org/viewarticle/569207 (accessed Jul. 03, 2023).

[132] M. Yanada, K. Harada, Y. Shimomura, Y. Arai, and T. Konuma, “Conditioning regimens for allogeneic hematopoietic cell transplantation in acute myeloid leukemia: Real-world data from the Japanese registry studies,” Frontiers in Oncology, vol. 12, 2022, Accessed: Jul. 04, 2023. [Online]. Available: https://www.frontiersin.org/articles/10.3389/fonc.2022.1050633

[133] F. Saraceni et al., “Conditioning Regimens for Frail Patients with Acute Leukemia Undergoing Allogeneic Stem Cell Transplant: How to Strike Gently,” Clin Hematol Int, vol. 3, no. 4, pp. 153–160, Aug. 2021, doi: 10.2991/chi.k.210731.001.

[134] Y. S. Jethava et al., “Conditioning regimens for allogeneic hematopoietic stem cell transplants in acute myeloid leukemia,” Bone Marrow Transplant, vol. 52, no. 11, Art. no. 11, Nov. 2017, doi: 10.1038/bmt.2017.83.

[135] F. Khimani et al., “Impact of Total Body Irradiation-Based Myeloablative Conditioning Regimens in Patients with Acute Lymphoblastic Leukemia Undergoing Allogeneic Hematopoietic Stem Cell Transplantation: Systematic Review and Meta-Analysis,” Transplant Cell Ther, vol. 27, no. 7, p. 620.e1-620.e9, Jul. 2021, doi: 10.1016/j.jtct.2021.03.026.

[136] C. Luo et al., “Myeloablative conditioning regimens in adult patients with acute myeloid leukemia undergoing allogeneic hematopoietic stem cell transplantation in complete remission: a systematic review and network meta-analysis,” Bone Marrow Transplant, vol. 58, no. 2, Art. no. 2, Feb. 2023, doi: 10.1038/s41409-022-01865-6.

[137] “Pre-Stem Cell Transplant Treatment Regimen for AML, MDS - NCI,” May 26, 2017. https://www.cancer.gov/news-events/cancer-currents-blog/2017/aml-stem-cell-transplant (accessed Jul. 04, 2023).

[138] A. Nagler and A. Shimoni, “Conditioning,” in The EBMT Handbook: Hematopoietic Stem Cell Transplantation and Cellular Therapies, E. Carreras, C. Dufour, M. Mohty, and N. Kröger, Eds., 7th ed.Cham (CH): Springer, 2019. Accessed: Jul. 04, 2023. [Online]. Available: http://www.ncbi.nlm.nih.gov/books/NBK553926/

[139] P. Venkatesh and A. Kasi, “Anthracyclines,” in StatPearls, Treasure Island (FL): StatPearls Publishing, 2023. Accessed: Jul. 04, 2023. [Online]. Available: http://www.ncbi.nlm.nih.gov/books/NBK538187/

[140] L. Law, J. Tuscano, T. Wun, K. Ahlberg, and C. Richman, “Filgrastim treatment of acute myelogenous leukemia (M7) relapse after allogeneic peripheral stem cell transplantation resulting in both graft-versus-leukemia effect with cytogenetic remission and chronic graft-versus-host disease manifesting as polyserositis and subsequent bronchiolitis obliterans with organizing pneumonia,” Int J Hematol, vol. 76, no. 4, pp. 360–364, Nov. 2002, doi: 10.1007/BF02982697.

[141] N. B. Granville, F. Rubio, A. Unugur, E. Schulman, and W. Dameshek, “Treatment of Acute Leukemia in Adults with Massive Doses of Prednisone and Prednisolone,” New England Journal of Medicine, vol. 259, no. 5, pp. 207–213, Jul. 1958, doi: 10.1056/NEJM195807312590502.

[142] H. Inaba and C.-H. Pui, “Glucocorticoid use in acute lymphoblastic leukemia: comparison of prednisone and dexamethasone,” Lancet Oncol, vol. 11, no. 11, pp. 1096– 1106, Nov. 2010, doi: 10.1016/S1470-2045(10)70114-5.

[143] T. Ono, “Which Tyrosine Kinase Inhibitors Should Be Selected as the First-Line Treatment for Chronic Myelogenous Leukemia in Chronic Phase?,” Cancers (Basel), vol. 13, no. 20, p. 5116, Oct. 2021, doi: 10.3390/cancers13205116.

[144] “Tyrosine kinase inhibitors (TKI) for CML | Macmillan Cancer Support.” https://www.macmillan.org.uk/cancer-information-and-support/treatments-and-drugs/tyrosine-kinase-inhibitors-for-chronic-myeloid-leukaemia (accessed Jul. 04, 2023).

[145] “Chronic Myelogenous Leukemia Treatment - NCI,” Jun. 30, 2023. https://www.cancer.gov/types/leukemia/patient/cml-treatment-pdq (accessed Jul. 04, 2023).

[146] C. for D. E. and Research, “FDA approves Onureg (azacitidine tablets) for acute myeloid leukemia,” FDA, Sep. 2020, Accessed: Jul. 04, 2023. [Online]. Available: https://www.fda.gov/drugs/resources-information-approved-drugs/fda-approves-onureg-azacitidine-tablets-acute-myeloid-leukemia

[147] A. C. Schuh, H. Döhner, L. Pleyer, J. F. Seymour, P. Fenaux, and H. Dombret, “Azacitidine in adult patients with acute myeloid leukemia,” Crit Rev Oncol Hematol, vol. 116, pp. 159–177, Aug. 2017, doi: 10.1016/j.critrevonc.2017.05.010.

[148] “Azacitidine - NCI,” Oct. 05, 2006. https://www.cancer.gov/about-cancer/treatment/drugs/azacitidine (accessed Jul. 04, 2023).

[149] F. Huguet et al., “Clofarabine for the treatment of adult acute lymphoid leukemia: the Group for Research on Adult Acute Lymphoblastic Leukemia intergroup,” Leuk Lymphoma, vol. 56, no. 4, pp. 847–857, Apr. 2015, doi: 10.3109/10428194.2014.887708.

[150] A. W. Rijneveld et al., “Clofarabine added to intensive treatment in adult patients with newly diagnosed ALL: the HOVON-100 trial,” Blood advances, vol. 6, no. 4, pp. 1115–1125, 2022.

[151] S. Nakako et al., “Successful management of therapy-related chronic myelomonocytic leukemia with cytarabine, aclarubicin, and azacitidine following tegafur/gimeracil/oteracil,” Clin Case Rep, vol. 9, no. 6, p. e04298, Jun. 2021, doi: 10.1002/ccr3.4298.

[152] D. Machover et al., “Phase I-II study of aclarubicin for treatment of acute myeloid leukemia,” Cancer Treat Rep, vol. 68, no. 6, pp. 881–886, Jun. 1984.

[153] A. Hochhaus and H. Kantarjian, “The development of dasatinib as a treatment for chronic myeloid leukemia (CML): from initial studies to application in newly diagnosed patients,” J Cancer Res Clin Oncol, vol. 139, no. 12, pp. 1971–1984, 2013, doi: 10.1007/s00432-013-1488-z.

[154] M. Talpaz, G. Saglio, E. Atallah, and P. Rousselot, “Dasatinib dose management for the treatment of chronic myeloid leukemia,” Cancer, vol. 124, no. 8, pp. 1660– 1672, Apr. 2018, doi: 10.1002/cncr.31232.

[155] R. Foà et al., “Dasatinib–blinatumomab for Ph-positive acute lymphoblastic leukemia in adults,” New England Journal of Medicine, vol. 383, no. 17, pp. 1613–1623, 2020.

[156] Z. T. Han et al., “Effect of intravenous infusions of 12-O-tetradecanoylphorbol-13-acetate (TPA) in patients with myelocytic leukemia: Preliminary studies on therapeutic efficacy and toxicity,” Proc. Natl. Acad. Sci. U.S.A., vol. 95, no. 9, pp. 5357–5361, Apr. 1998, doi: 10.1073/pnas.95.9.5357.

[157] X. Zheng et al., “Synergistic stimulatory effect of 12-O-tetradecanoylphorbol-13-acetate and capsaicin on macrophage differentiation in HL-60 and HL-525 human myeloid leukemia cells,” Int J Oncol, vol. 26, no. 2, pp. 441–448, Feb. 2005.

[158] X. Zheng et al., “Gene expression of TPA induced differentiation in HL-60 cells by DNA microarray analysis,” Nucleic Acids Res, vol. 30, no. 20, pp. 4489– 4499, Oct. 2002, doi: 10.1093/nar/gkf580.

[159] W. G. Wierda and F. P. Tambaro, “How I manage CLL with venetoclax-based treatments,” Blood, The Journal of the American Society of Hematology, vol. 135, no. 17, pp. 1421–1427, 2020.

[160] J. Jumper et al., “Highly accurate protein structure prediction with AlphaFold,” Nature, vol. 596, no. 7873, Art. no. 7873, Aug. 2021, doi: 10.1038/s41586-021-03819-2.

[161] A. A. Kiaei and H. Khotanlou, “Segmentation of medical images using mean value guided contour,” Medical Image Analysis, vol. 40, pp. 111–132, Aug. 2017, doi: 10.1016/j.media.2017.06.005.

[162] R. Bahadori et al., “Diagnosing Alzheimer’s Disease Levels Using Machine Learning and MRI: A Novel Approach.” Preprints, Jun. 16, 2023. doi: 10.20944/preprints202306.1184.v1.

[163] H. Jafari et al., “A full pipeline of diagnosis and prognosis the risk of chronic diseases using deep learning and Shapley values: The Ravansar county anthropometric cohort study,” PLOS ONE, vol. 17, no. 1, p. e0262701, Jan. 2022, doi: 10.1371/journal.pone.0262701.

[164] A. A. Kiaei et al., “Active Identity Function as Activation Functio.” 2023. doi: 10.20944/preprints202305.1018.v1.

[165] N. Salari et al., “Executive protocol designed for new review study called: systematic review and artificial intelligence network meta-analysis (RAIN) with the first application for COVID-19,” Biology Methods and Protocols, 2023.

[166] M. Boush, A. A. Kiaei, D. Safaei, S. Abadijou, N. Salari, and M. Mohammadi, “Drug combinations proposed by machine learning on genes/proteins to improve the efficacy of Tecovirimat in the treatment of Monkeypox: A Systematic Review and Network Meta-analysis.” medRxiv, p. 2023.04.23.23289008, Apr. 25, 2023. doi: 10.1101/2023.04.23.23289008.

[167] A. Kiaei et al., “Identification of suitable drug combinations for treating COVID-19 using a novel machine learning approach: The RAIN method,” Life, vol. 12, no. 9, p. 1456, 2022.

[168] “Recommending Drug Combinations Using Reinforcement Learning targeting Genes/proteins associated with Heterozygous Familial Hypercholesterolemia: A comprehensive Systematic Review and Net-work Meta-analysis,” Feb. 16, 2023. https://www.researchsquare.com (accessed Jul. 04, 2023).

[169] M. Boush, A. A. Kiaei, D. Safaei, S. Abadijou, N. Salari, and M. Mohammadi, “Recommending Drug Combinations using Reinforcement Learning to target Genes/proteins that cause Stroke: A comprehensive Systematic Review and Network Meta-analysis.” medRxiv, p. 2023.04.20.23288906, Apr. 24, 2023. doi: 10.1101/2023.04.20.23288906.

[170] D. Safaei et al., “Systematic review and network metaanalysis of drug combinations suggested by machine learning on genes and proteins, with the aim of improving the effectiveness of Ipilimumab in treating Melanoma.” medRxiv, p. 2023.05.13.23289940, May 25, 2023. doi: 10.1101/2023.05.13.23289940.

[171] L. Heraudet et al., “VANDA regimen followed by blinatumomab leads to favourable outcome in patients with Philadelphia chromosome-negative B-precursor acute lymphoblastic leukaemia in first relapse,” Br J Haematol, vol. 198, no. 3, pp. 523–527, Aug. 2022, doi: 10.1111/bjh.18218.

[172] K. Moriya et al., “The incidence of symptomatic osteonecrosis is similar between Japanese children and children in Western countries with acute lymphoblastic leukaemia treated with a Berlin-Frankfurt-Münster (BFM)95-based protocol,” Br J Haematol, vol. 196, no. 5, pp. 1257–1261, Mar. 2022, doi: 10.1111/bjh.17988.

[173] A. Iannuzzi et al., “Case Report: Genetic Analysis of PEG-Asparaginase Induced Severe Hypertriglyceridemia in an Adult With Acute Lymphoblastic Leukaemia,” Front Genet, vol. 13, p. 832890, 2022, doi: 10.3389/fgene.2022.832890.

[174] B. Yohannan, F. Cervoni-Curet, and A. Rios, “AML-173 Pseudotumor Cerebri (PTC) in Patients With Acute Leukemia (AL),” Clinical Lymphoma Myeloma and Leukemia, vol. 22, pp. S221–S222, Oct. 2022, doi: 10.1016/S2152-2650(22)01237-X.

[175] C. Canter et al., “AML-434 Retrospective Review of Outcomes in Newly Diagnosed Leukemia Patients Aged 65 and Older When Admitted to the Hospital: A Single Center Experience,” Clinical Lymphoma Myeloma and Leukemia, vol. 22, p. S250, Oct. 2022, doi: 10.1016/S2152-2650(22)01294-0.

[176] L. Pardo Gambarte et al., “ZBTB16-RARα-Positive Atypical Promyelocytic Leukemia: A Case Report,” Medicina (Kaunas), vol. 58, no. 4, p. 520, Apr. 2022, doi: 10.3390/medicina58040520.

[177] H. M. Kantarjian et al., “The cure of leukemia through the optimist’s prism,” Cancer, vol. 128, no. 2, pp. 240–259, Jan. 2022, doi: 10.1002/cncr.33933.

[178] S. Takahashi, “Kinase Inhibitors and Interferons as Other Myeloid Differentiation Inducers in Leukemia Therapy,” Acta Haematol, vol. 145, no. 2, pp. 113–121, 2022, doi: 10.1159/000519769.

[179] J. Zhao et al., “Case report: A rare case of acute myeloid leukemia with CPSF6-RARG fusion resembling acute promyelocytic leukemia,” Front Oncol, vol. 12, p. 1011023, 2022, doi: 10.3389/fonc.2022.1011023.

[180] N. Lin, Z. Liu, Y. Li, X. Yan, and L. Wang, “Determining the Appropriate Treatment for T-Cell Acute Lymphoblastic Leukemia With SET-CAN/NUP214 Fusion: Perspectives From a Case Report and Literature Review,” Front Oncol, vol. 11, p. 651494, 2021, doi: 10.3389/fonc.2021.651494.

[181] B. L. Z. Oh et al., “Successful toxicity reduction during delayed intensification in the non-high-risk arm of Malaysia-Singapore Acute Lymphoblastic Leukaemia 2010 study,” Eur J Cancer, vol. 142, pp. 92–101, Jan. 2021, doi: 10.1016/j.ejca.2020.10.010.

[182] N. Jabbar, N. Khayyam, U. Arshad, S. Maqsood, S. A. Hamid, and N. Mansoor, “An Outcome Analysis of Childhood Acute Promyelocytic Leukemia Treated with Atra and Arsenic Trioxide, and Limited Dose Anthracycline,” Indian J Hematol Blood Transfus, vol. 37, no. 4, pp. 569–575, Oct. 2021, doi: 10.1007/s12288-021-01404-1.

[183] P. Schneider et al., “Decitabine mildly attenuates MLL-rearranged acute lymphoblastic leukemia in vivo, and represents a poor chemo-sensitizer,” EJHaem, vol. 1, no. 2, pp. 527–536, Nov. 2020, doi: 10.1002/jha2.81.

[184] Y. Sun et al., “Transformation from acute promyelocytic leukemia to acute myeloid leukemia with a CEBPA double mutation: A case report and review of the literature,” Medicine (Baltimore), vol. 100, no. 5, p. e24385, Feb. 2021, doi: 10.1097/MD.0000000000024385.

[185] J. I. Traylor et al., “Computational Drug Repositioning Identifies Potentially Active Therapies for Chordoma,” Neurosurgery, vol. 88, no. 2, pp. 428–436, Jan. 2021, doi: 10.1093/neuros/nyaa398.

[186] S. Takahashi, “Current Understandings of Myeloid Differentiation Inducers in Leukemia Therapy,” Acta Haematol, vol. 144, no. 4, pp. 380–388, 2021, doi: 10.1159/000510980.

[187] E. Jabbour et al., “Hyper-CVAD regimen in combination with ofatumumab as frontline therapy for adults with Philadelphia chromosome-negative B-cell acute lymphoblastic leukaemia: a single-arm, phase 2 trial,” Lancet Haematol, vol. 7, no. 7, pp. e523–e533, Jul. 2020, doi: 10.1016/S2352-3026(20)30144-7.

[188] A. Trobisch et al., “Invasive mucormycosis during treatment for acute lymphoblastic leukaemia-successful management of two life-threatening diseases,” Support Care Cancer, vol. 28, no. 5, pp. 2157–2161, May 2020, doi: 10.1007/s00520-019-04962-3.

[189] F. Pedrosa et al., “Reduced-dose intensity therapy for pediatric lymphoblastic leukemia: long-term results of the Recife RELLA05 pilot study,” Blood, vol. 135, no. 17, pp. 1458–1466, Apr. 2020, doi: 10.1182/blood.2019004215.

[190] M. A. Kutny et al., “Outcome for pediatric acute promyelocytic leukemia patients at Children’s Oncology Group sites on the Leukemia Intergroup Study CALGB 9710 (Alliance),” Pediatr Blood Cancer, vol. 66, no. 3, p. e27542, Mar. 2019, doi: 10.1002/pbc.27542.

[191] Y. Le, H. Li, C. Wan, Y. Long, and Z. Liu, “Rhabdomyolysis during myelosuppression in a patient with central nervous system leukemia: A case report,” Medicine (Baltimore), vol. 97, no. 45, p. e13091, Nov. 2018, doi: 10.1097/MD.0000000000013091.

[192] C. Wu and W. Li, “Genomics and pharmacogenomics of pediatric acute lymphoblastic leukemia,” Crit Rev Oncol Hematol, vol. 126, pp. 100–111, Jun. 2018, doi: 10.1016/j.critrevonc.2018.04.002.

[193] W. L. Salzer et al., “Toxicity associated with intensive postinduction therapy incorporating clofarabine in the very high-risk stratum of patients with newly diagnosed high-risk B-lymphoblastic leukemia: A report from the Children’s Oncology Group study AALL1131,” Cancer, vol. 124, no. 6, pp. 1150–1159, Mar. 2018, doi: 10.1002/cncr.31099.

[194] N. S. Araújo et al., “A Rare Case of Relapsed Pediatric Acute Promyelocytic Leukemia with Skin Involvement by Myeloid Sarcoma,” Am J Case Rep, vol. 19, pp. 438–441, Apr. 2018, doi: 10.12659/ajcr.907847.

[195] “The global problem of early deaths in acute promyelocytic leukemia: A strategy to decrease induction mortality in the most curable leukemia - PubMed.” https://pubmed.ncbi.nlm.nih.gov/29033137/ (accessed Jul. 08, 2023).

[196] N. S. Fracchiolla, M. Sciumè, G. Cernuschi, and A. Cortelezzi, “An Unusual Coexistence of Primary Central Nervous System Non-Hodgkin’s Lymphoma and Acute Promyelocytic Leukemia,” Case Rep Hematol, vol. 2018, p. 2741939, 2018, doi: 10.1155/2018/2741939.

[197] B. G. Mar et al., “SETD2 alterations impair DNA damage recognition and lead to resistance to chemotherapy in leukemia,” Blood, vol. 130, no. 24, pp. 2631–2641, Dec. 2017, doi: 10.1182/blood-2017-03-775569.

[198] K. Teiken, H. Kreipe, B. Maecker-Kolhoff, and K. Hussein, “Variant of classical high grade PTLD: posttransplant EBV-negative T cell lymphoblastic leukaemia after solid organ transplantation,” Ann Hematol, vol. 96, no. 8, pp. 1403–1405, Aug. 2017, doi: 10.1007/s00277-017-3026-6.

[199] K. Horibe et al., “Long-term Results of the Risk-adapted Treatment for Childhood B-Cell Acute Lymphoblastic Leukemia: Report From the Japan Association of Childhood Leukemia Study ALL-97 Trial,” J Pediatr Hematol Oncol, vol. 39, no. 2, pp. 81–89, Mar. 2017, doi: 10.1097/MPH.0000000000000760.

[200] M. Song, X. Wu, M. Zhang, and P. He, “[Detection and clinical significance of PML protein expression of acute promyelocytic leukemia cells],” Xi Bao Yu Fen Zi Mian Yi Xue Za Zhi, vol. 33, no. 7, pp. 971–976, Jul. 2017.

[201] J. M. Brandwein et al., “Treatment of older patients with acute myeloid leukemia (AML): revised Canadian consensus guidelines,” Am J Blood Res, vol. 7, no. 4, pp. 30–40, 2017.

[202] J. H. Song, S. H. Kim, K.-M. Cho, S. Y. Hwang, H.-J. Kim, and T. S. Kim, “Analysis of gene profiles involved in the enhancement of all-trans retinoic acid-induced HL-60 cell differentiation by sesquiterpene lactones identifies asparagine synthetase as a novel target for differentiation-inducing therapy,” Int J Oncol, vol. 44, no. 3, pp. 970–976, Mar. 2014, doi: 10.3892/ijo.2013.2241.

[203] R. Castelli, B. Ferrari, A. Cortelezzi, and A. Guariglia, “Thromboembolic complications in malignant haematological disorders,” Curr Vasc Pharmacol, vol. 8, no. 4, pp. 482–494, Jul. 2010, doi: 10.2174/157016110791330799.

[204] S. J. Corey, “New agents in the treatment of childhood leukemias and myelodysplastic syndromes,” Curr Oncol Rep, vol. 7, no. 6, pp. 399–405, Nov. 2005, doi: 10.1007/s11912-005-0003-3.

[205] C. Langebrake, D. Reinhardt, and J. Ritter, “Minimising the long-term adverse effects of childhood leukaemia therapy,” Drug Saf, vol. 25, no. 15, pp. 1057–1077, 2002, doi: 10.2165/00002018-200225150-00002.

